# Introduction and community transmission of SARS-CoV-2 lineage A.2.5 in Florida with novel spike INDELS

**DOI:** 10.1101/2021.12.03.21266538

**Authors:** Sarah E. Schmedes, Taj Azarian, Eleonora Cella, Jessy Motes, Omer Tekin, James Weiss, Nancimae Miller, Jason Blanton

## Abstract

SARS-CoV-2 (SC2) variants of concern (VOC) continue to emerge and spread globally, threatening the use of monoclonal antibody therapies and vaccine effectiveness. Several mutations in the SC2 spike glycoprotein have been associated with reduction in antibody neutralization. Genomic surveillance of SC2 variants has been imperative to inform the public health response regarding the use of clinical therapies in specific jurisdictions based on the proportion of particular variants (e.g., Gamma (P.1)) in a region. Florida Department of Health Bureau of Public Health Laboratories (BPHL) performs tiled-amplicon whole genome sequencing for baseline and targeted surveillance of SC2 isolates in Florida from clinical specimens collected from county health departments and hospitals throughout the state. Here, we describe the introduction of SC2 lineage A.2.5 in Florida, which contains S:L452R (a substitution of therapeutic concern) and two novel Spike INDELS, the deletion of 141-143 and ins215AGY, with unknown implications on immune response. The A.2.5 lineage was first detected in Florida among an outbreak at a healthcare facility in January 2021, and subsequent A.2.5 isolates were detected across all geographical regions throughout the state. A time-scaled maximum clade credibility phylogeny determined there were at least eight separate introductions of A.2.5 in the state. The time of introduction of a monophyletic Florida clade was established to be December 2020. The Spike INDELS were determined to reside in the N-terminal domain, a region associated with antibody neutralization. As community transmission of SARS-CoV-2 in Florida continues, genomic surveillance of circulating variants in Florida and the detection of emerging variants are critical for informing public health response to COVID-19.

## Introduction

SARS-CoV-2 (SC2) variants of concern continue to emerge and spread globally, highlighting the demand for increased SC2 surveillance at the local, national, and international level. Tiled-amplicon whole-genome sequencing (WGS) or targeted enrichment WGS has been the gold standard for SC2 surveillance to identify and track lineages which contain amino acid substitutions and/or insertions/deletions (INDELS) within the spike glycoprotein, which binds to the ACE-2 receptor during infection. Several spike glycoprotein mutations in VOCs have shown to have significant public health impacts such as increased transmission (1-4) and may pose a threat to front-line therapeutics and vaccines (4-9). Active SC2 surveillance by WGS is imperative to detect novel spike mutations as soon as they emerge.

The Centers for Disease Control and Prevention (CDC) published a United States (U.S.) government interagency variant classification list characterizing variants in the United States with defined attributes: variant of interest (VOI), variant of concern (VOC), and variant of high consequence (VOHC) (10). As of June 4, 2021, VOCs and VOIs comprised the majority of all lineages in the U.S. Among U.S. jurisdictions, Florida had one of the highest burdens of VOCs (> 80%) amid lineages circulating within the state (11).

Florida Department of Health (FDOH) Bureau of Public Health Laboratories (BPHL) has sequenced COVID-19-positive samples collected from patients throughout the state, as early as March 4, 2020. BPHL actively surveils lineages of public health interest in Florida, including variant classifications designated by CDC, WHO, and other international organizations (10, 12, 13), and actively monitors changes in lineage frequencies in the state in addition to the emergence of novel spike mutations which may warrant further investigation.

In January 2021, BPHL detected the first cases of then SC2 lineage A.2.4 among an outbreak at a south Florida healthcare facility. Shortly after this first detection, additional A.2.4 cases were detected in routine surveillance samples collected from patients geographically dispersed across the state. The A.2.4 lineage, which first emerged in Panama (14), comprised a distinct clade which contained mainly U.S. samples, including those from Florida. This separate clade was soon reclassified as lineage A.2.5 and was defined as a Central American/U.S. lineage (15).

SC2 A.2.5 isolates detected in Florida became of particular interest as this lineage was first detected among a healthcare facility outbreak involving both patients and staff. The A.2.5 isolates contained several spike mutations of concern which warranted further investigation, including a CDC-designated substitution of therapeutic concern (L452R), the independent emergence of D614G in an A lineage, and two novel spike INDELs (LGV141-143del and ins215AGY) not yet seen in Florida, nor described in the literature. Here, we describe the initial introductions and spread of A.2.5 in Florida and further characterize spike glycoprotein mutations LGV141-143del and ins215AGY.

## Materials and Methods

### RNA extraction from clinical specimens

Primary specimens were received at BPHL and confirmed for the presence of SARS-CoV-2 RNA. RNA was purified by multiple methods, including: Kingfisher combined with the Thermo Fisher MagMAX Viral/Pathogen II (MVP II) Nucleic Acid Isolation kit (Applied Biosystems by Thermo Fisher Scientific, Waltham, MA); Abbott m2000 System combined with the Abbott RealTime SARS-CoV-2 assay (Abbott, Lake County, IL;, MagNA Pure 96 combined with the MagNA Pure 96 DNA and Viral NA Small Volume Kit (Roche, Indianapolis, IN); MagNA Pure LC 2.0 combined with the MagNA Pure LC Total Nucleic Acid Isolation Kit (Roche, Indianapolis, IN); and EZ1 Advanced combined with the EZ1 DSP Virus Kit (Qiagen, Germantown, MD), QIAamp DSP Viral RNA Mini Kit (Qiagen, Germantown, MD).

### cDNA and amplicon generation/Library preparation

The BPHL SARS-CoV-2 Tiled Amplicon Illumina Sequencing protocol (16) was followed.

### Sequencing

Libraries were sequenced on either an Illumina MiSeq or NextSeq 550 Dx. For the MiSeq: The protocol described at (16) was followed. For the NextSeq 550Dx: The sample pools were diluted to 4 nM based on the Qubit measurements and Agilent sizing information, and 5 µL of the 4 nM pool was denatured with 5 µL of 0.2 N NaOH. Amplicon libraries were diluted to 1.5 pM in Illumina’s HT1 buffer, spiked with 1% PhiX, and sequenced using a NextSeq 500/550 Mid Output Kit v2.5 (300 cycle).

### Consensus assembly generation and variant profiles

SARS-CoV-2 consensus assemblies were generated using a custom pipeline, FLAQ-SC2. Quality metrics were generated for raw sequence reads using fastqc v0.11.9 (http://www.bioinformatics.babraham.ac.uk/projects/fastqc/), and raw fastqs were quality filtered and trimmed (SLIDINGWINDOW:4:30 MINLEN:75 TRAILING:20) using trimmomatic v0.39 (17). Sequence adapter and Phix contamination were removed using bbduk (BBMap v38.79) (https://sourceforge.net/projects/bbmap/). Cleaned reads were mapped to SARS-CoV-2 reference NC_045512.2 using bwa mem v0.7.17-r1188 (18). PCR duplicates were marked and removed using samtools v1.9 (19). ARTIC primer sequences (https://github.com/artic-network/artic-ncov2019/tree/master/primer_schemes/nCoV-2019/V3) were trimmed from bam files using ivar trim v1.2.2 (20). Variants were called using samtools mpileup v1.9 (19) and ivar variant v1.2.2 (20), and consensus assemblies were generated using samtools mpileup v1.9 (19) and ivar consensus v1.2.2 (20). Only variants as the major allele were included in the final consensus assembly, and bases <10x were incorporated as Ns. Read mapping metrics were generated using samtools coverage v1.10 (19). Only samples with ≥80% genome coverage at ≥100x mean read depth were submitted to public repositories (i.e., NCBI Genbank (21) and GISAID (https://www.gisaid.org/)) and included in this study. SC2 lineages (22) were determined using Pangolin v2.3.6-pangolearn-2021-03-16 (https://github.com/cov-lineages/pangolin). Amino acid substitution and INDEL data were also generated using the command-line version of NextClade v0.14.1 (https://clades.nextstrain.org/).

### Data availability

BPHL A.2.5 consensus genomes are available on NCBI Genbank (MW586581-MW586584, MW586586, MW586589, MW689883, MW689890, MW689894, MW689907, MW715557, MW725772, MW735564, MW735565, MW737635, MW742685, MW810328, MW810330) and GISAID (EPI_ISL_965016-EPI_ISL_965019, EPI_ISL_965021, EPI_ISL_965024, EPI_ISL_1132181, EPI_ISL_1132278-EPI_ISL_1132280, EPI_ISL_1191584, EPI_ISL_1218839, EPI_ISL_1238766, EPI_ISL_1239972, EPI_ISL_1239973, EPI_ISL_1250852, EPI_ISL_1372280, EPI_ISL_1372282).

### Public SC2 data

Published genomes and associated metadata for all global A.2.5 isolates (n=220) were downloaded from GISAID (https://www.gisaid.org/) on 2021-03-29 (with “complete” and “low coverage excluded” checked). Thirty-three A.2.5 isolates were from Florida, including eighteen isolates sequenced at BPHL. Pangolin v2.3.6-pangolearn-2021-03-16 (https://github.com/cov-lineages/pangolin) and NextClade v0.14.1 (https://clades.nextstrain.org/) were run on all data to confirm lineage and variants as included in the GISAID sample metadata.

### Phylogenetic and coalescent analysis

Full genome sequences were aligned with MAFFT v7.475 (23), and a maximum likelihood phylogeny was inferred using IQtree (24) with the following settings: iqtree -T AUTO -s aligned.fasta -m MFP -nt AUTO - B 1000. TreeTime was used to assess the temporal signal by performing a linear regression of the root-to-tip genetic distances against sampling dates (25). A coalescent analysis was carried out using BEAST v1.10.4 (26) with a strict molecular clock model, HKY nucleotide substitution model, and exponential growth coalescent model. Triplicate Markov chain Monte Carlo (MCMC) runs of 100 million states were computed, sampling every 10,000 steps. Convergence of MCMC chains was checked using Tracer v.1.7.1 (27). A maximum clade credibility (MCC) tree was summarized from the posterior distribution of trees using TreeAnnotator with a 10% burn-in.

### Spike protein structural characterization

The spike glycoprotein sequence from strain hCoV-19/USA/FL-BPHL-0057/2021 (MW586582; EPI_ISL_965017) was used to model the three-dimensional structure using SWISS-MODEL (https://swissmodel.expasy.org/) with template SMTL ID 7cn8.1, which was identified as the closest match to the A.2.5 target sequence (28). The structure was then visualized and annotated with PyMOL v2.4.2 (29).

### Ethics Statement

This work has been reviewed by the Florida Department of Health Ethics and Human Research Protection Program and the Institutional Review Board determined that the ‘activity is public health practice and not research involving human subjects.’

## Results

On March 29, 2021, the global A.2.5 lineage comprised 220 isolates with complete genomes from 13 countries (Figure 1A). The earliest collected A.2.5 clinical isolate submitted to GISAID was collected in Ecuador on December 1, 2020. Thirty-three A.2.5 isolates were from Florida, and the first clinical specimen in Florida was collected on January 16, 2021. Assessment of the S gene revealed that the most common spike protein mutations included S:D614G (98.64%), S:L452R (97.27%), S:G142-(97.27%), S:V143- (97.27%), S:L141- (95.45%), S:D215A (76.82%), and S:ins215AGY (76.82%). Samples from FL depicted similar mutations frequencies S:D614G (96.97%), S:L452R (96.97%), S:G142- (100%), S:V143- (100%), S:L141- (100%), S:D215A (87.88%), and S:ins215AGY (87.88%).

**Figure 1.**
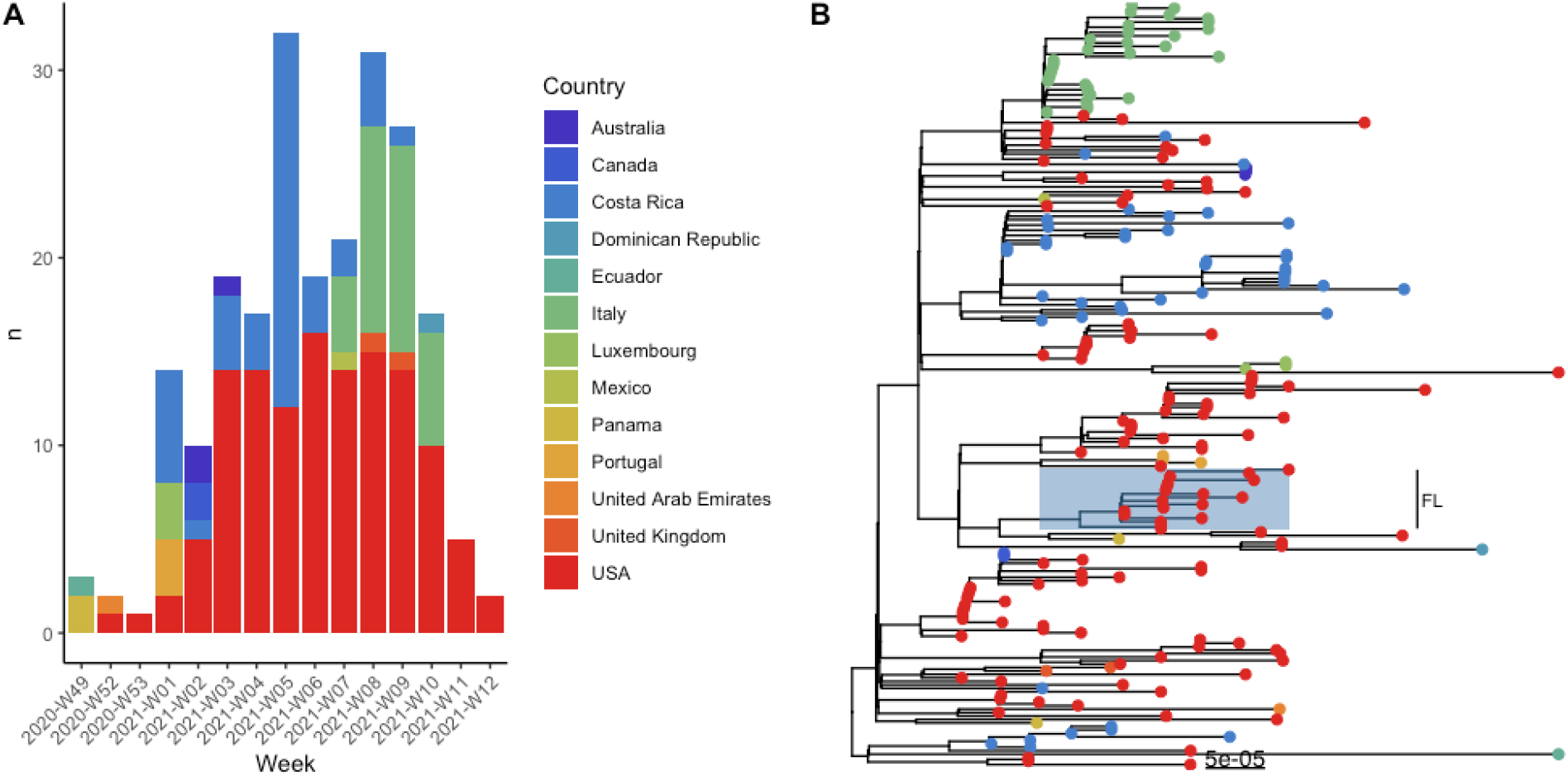
Time series of SARS-CoV-2 lineage A.2.5 isolates by collection date and country of isolation (A) and maximum likelihood phylogeny of 220 A.2.5 global isolates (B). Bars in (A) are colored by country of isolation and correspond to the tip point coloring in (B). The Florida clade, comprised of 17 isolates, is highlighted in light blue and annotated on the phylogeny.

### Introduction and continued community transmission of A.2.5 in Florida

A maximum likelihood phylogeny demonstrated that lineage A.2.5 is globally distributed, with dominant clades found in Central and South America as well as the United States and Italy (Figure 1B). The phylogeny also resolved a clade of 17 Florida isolates. The remaining isolates from Florida were interspersed throughout the phylogeny suggesting multiple importations of this emerging lineage.

To further investigate the emergence of lineage A.2.5, we performed a coalescent analysis. The maximum likelihood phylogeny (Figure 1B) was transformed into a time-scaled tree. It was then determined that sequence hCoV-19/Ecuador/USFQ-556/2020 was an outlier and was removed from the analysis. Assessment of temporal signal (i.e., molecular clock) indicated that the SARS-CoV-2 A.2.5 sequences evolved in a sufficient clock-like manner (r=0.26) (Figure 2A-B). Time-scaled analysis estimated the SARS-CoV-2 lineage A.2.5 evolutionary rate as 5.15E-4 substitutions/site/year (95% HPD, 4.07E-4 - 6.23E-4), consistent with the previous estimates (30).

**Figure 2.**
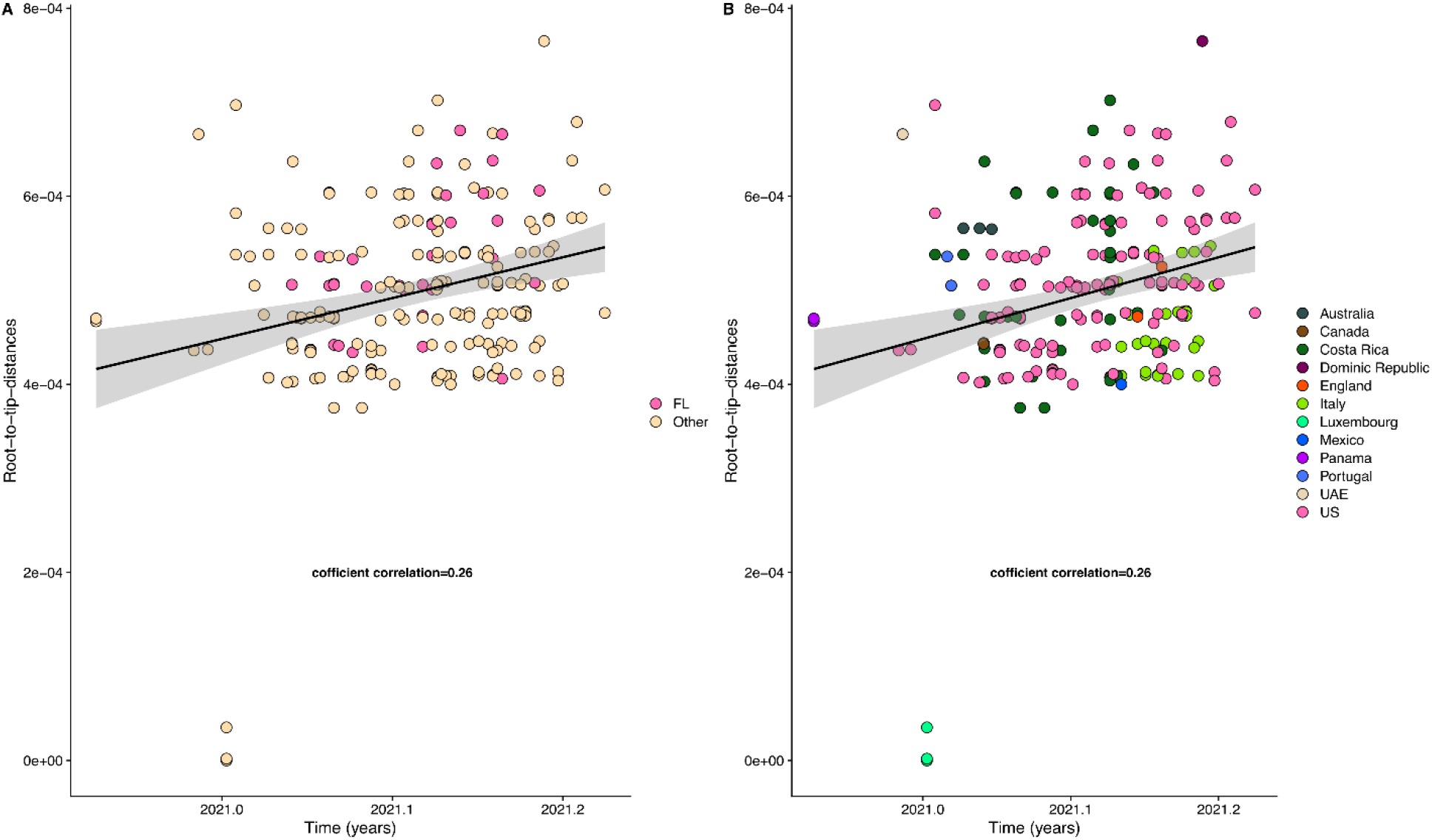
Temporal signal analysis of SARS-CoV-2 lineage A.2.5. For SARS-CoV-2, a temporal signal with correlation coefficient > 0.2 is generally accepted as sufficient temporal signal for coalescent analysis. The root-to-tip genetic distance is regressed on the collection date. Points are colored by U.S. or non-U.S. (A) or by country (B). Regression lines are shown with upper and lower error intervals (shaded grey area) representing 90% confidence intervals.

The most recent common ancestor (TMRCA) of the root ranged between late December 2019 and February 2020 (mean January 2020) (Figure 3). Strains from Luxembourg and one from the United States are located at the base of the main clade suggesting two independent and contemporaneous emergences of this lineage originated in the second half of 2020, when this lineage was first observed. The monophyletic clade comprising Florida strains originated in November 29, 2020 (95% HPD, October 24, 2020 – December 30, 2020); however, this early date was driven by two basal strains that may in fact represent a separate Florida introduction. Excluding those strains, the TMRCA of the clade is December 12, 2020 (95% HPD, November 27, 2020 – January 11, 2021). In addition to the dominant clade containing 17 strains, Florida isolates are interspersed throughout the maximum clade credibility (MCC) tree suggesting at least 8 separate introductions to the state (Figure 3).

**Figure 3.**
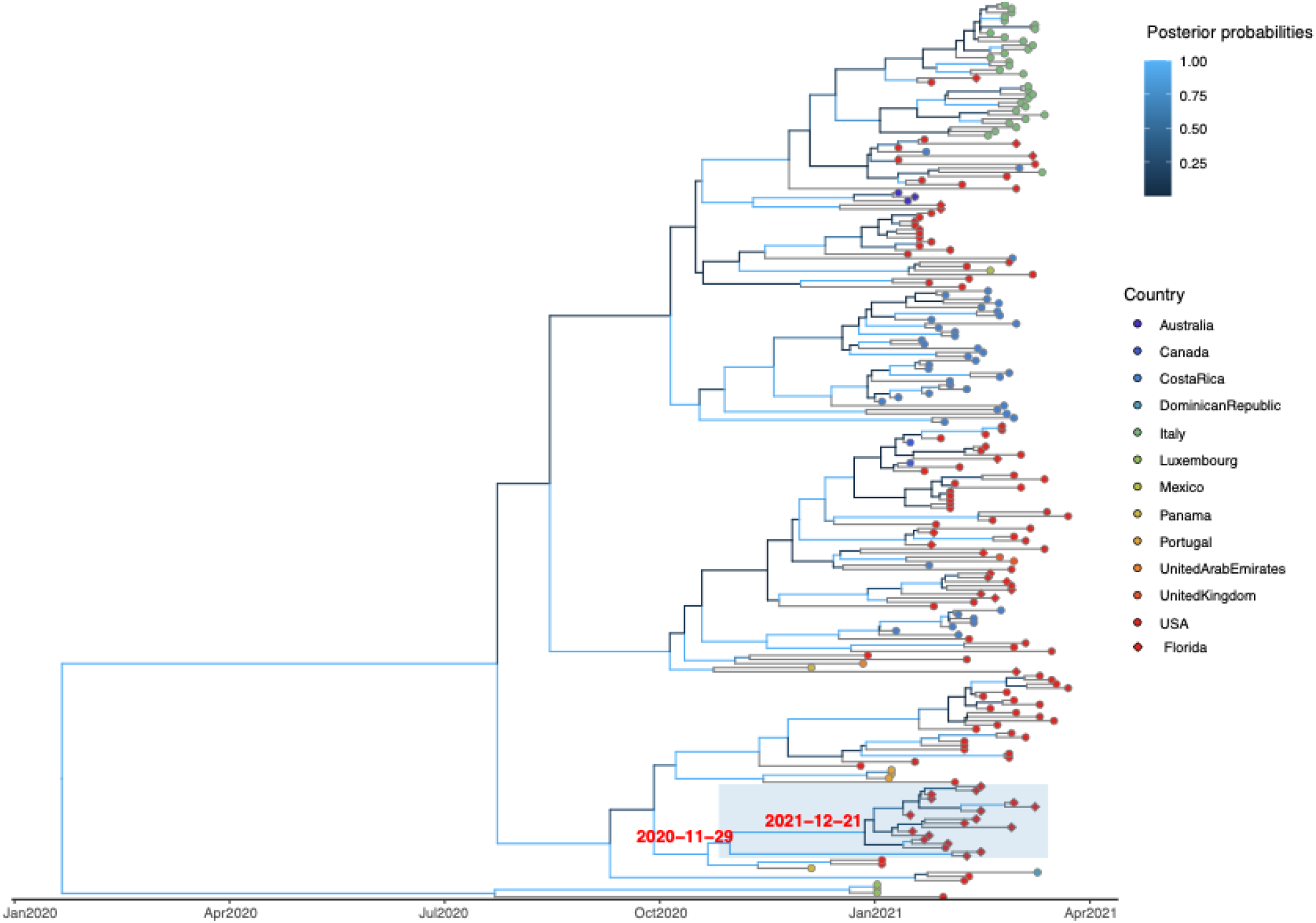
Maximum clade credibility tree of 219 SARS-CoV-2 lineage A.2.5 isolates. Branches are scaled in time and colored based on posterior probability. In general, a posterior probability above 0.8 is considered statistically significant. Tips are colored by country of isolation and isolates collected in Florida are represented by a diamond tip shape. The dominant Florida clade comprising 17 strains is highlighted, and the dates of the most recent common ancestor are labeled on the node.

### Three-dimensional spike glycoprotein structure with 141-143del and ins215AGY

The primary Florida clade all possessed Spike L141del, G142del, V143del, D215A, ins215AGY, L452R, and D614G, although 3/17 isolates had missing and low coverage at Spike 215. The deletions of Spike 141-143 and the insertion at 215 were most notable; therefore, we further explored the location of these INDELs by modeling their location on the three-dimensional structure of the spike glycoprotein. This modeling revealed that the INDELs both occurred in the N-terminal domain (NTD) (Figure 4).

**Figure 4.**
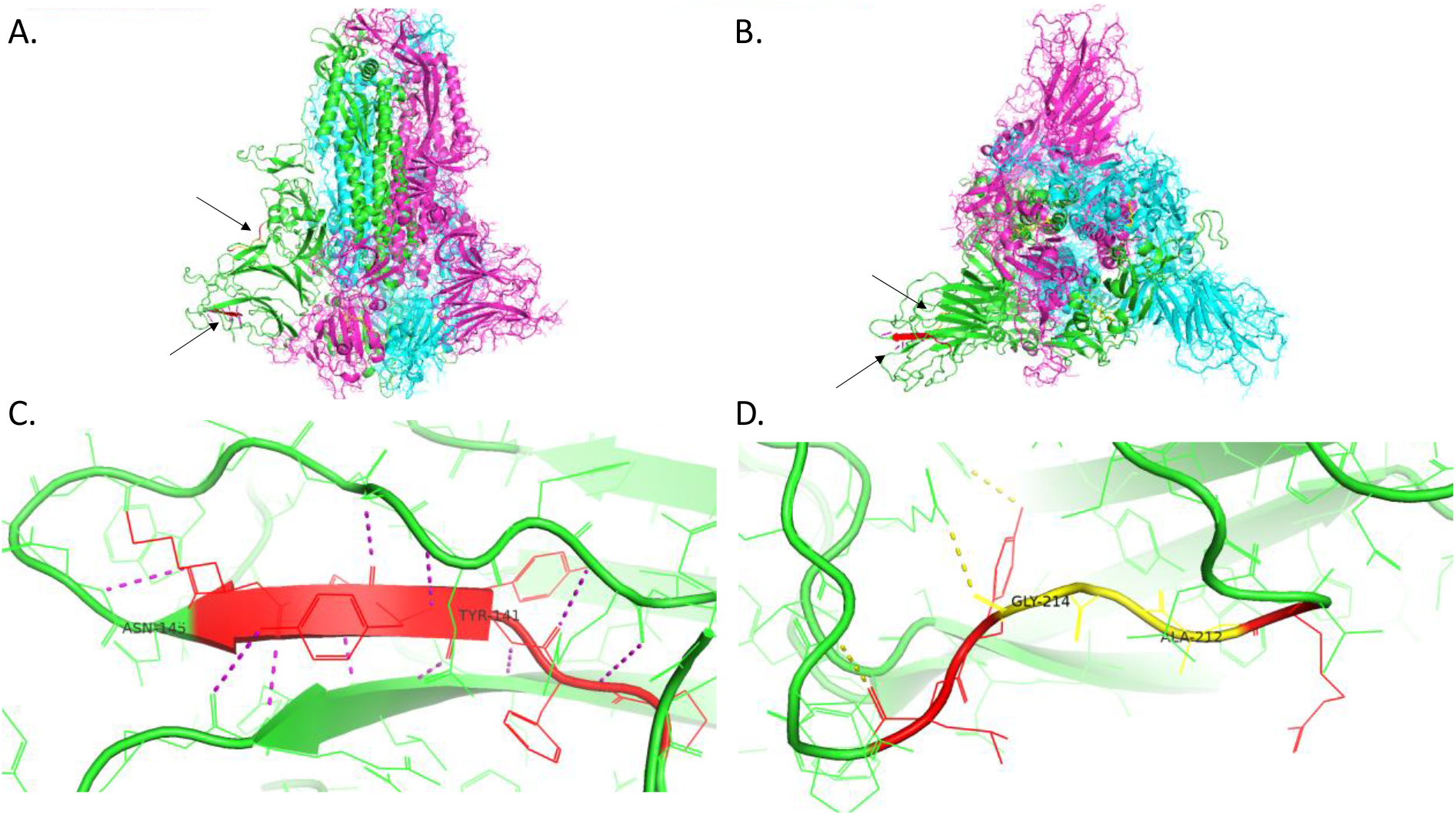
Three-dimensional structure of the SARS-CoV-2 spike glycoprotein illustrating the location of the A.2.5 INDELS 141-143 deletion and 215AGY insertion in the N-terminal domain. The structure is colored by domain and shown from the front view (A) and rotated to view the receptor binding site (B). The arrows point to the LGV deletion at Spike 141 and AGY insertion at 215, colored red to highlight their location in the NTD. The structural impact of the Spike 141-143 deletion and 215 insertion are shown in (C) and (D), respectively. Three flanking amino acids of the INDELs are colored in red, and insertions are colored yellow.

## Discussion

Since late 2020, SARS-CoV-2 variants continue to cause concern in the United States and globally. Spike protein mutations of therapeutic concern continue to emerge independently in lineages not classified as VOI or VOC (10), thus emphasizing the importance of SC2 genomic surveillance to detect novel substitutions of therapeutic concern (SOTC) as they emerge. Here, we described the introduction and transmission of lineage A.2.5 in Florida, which was introduced into the state near the end of 2020 around the same time as other VOCs first emerged in the U.S.

The A.2.5 lineage first emerged in Central America and the U.S. (15), and molecular clock analysis estimates the emergence of TMRCA of the root of the lineage at the beginning of the pandemic, late December 2019 – February 2020, with an introduction into Florida a year later around December 2020 (Figure 3). Lineage A.2.5 subsequently expanded rapidly to other regions around the world. In January 2020, A.2.5 specimens associated with an outbreak among patients and staff at a healthcare facility in Florida were detected, and subsequently after, sequenced A.2.5 specimens collected across the state from separate geographical locations, suggesting separate introductions of A.2.5 in Florida around the same time period, as supported by the MCC phylogeny (Figure 3).

The A.2.5 lineage in Florida became of particular interest due to the presence of mutations in the spike protein, associated with VOCs, including convergent evolution of D614G, a defining mutation among B.1 Pango lineages (31, 32), initially found in clades predominantly in Europe (33, 34). Spike D614G has become the dominant allele in isolates world-wide (34), and has been associated with increased viral loads (32, 34), infections among younger patients (32) and increased entry into human cells (35, 36), suggesting increased infectivity. A.2.5 also contains spike L452R, a CDC-defined SOTC (10), predominantly observed in Epsilon (B.1.427/B.1.429), which emerged in southern California in May 2020 and exhibits 20% increased transmissibility (4). Allele L452R has independently emerged in several other lineages over the last several months, including Iota (B.1.526), Kappa (B.1.617.1), Delta (B.1.617.2), and C.36 (10). Spike L452R causes increased infectivity (4) and has been reported to have a 15-fold reduction in susceptibility to combination monoclonal antibody therapy, bamlanivimab together with etesevimab (5).

Most notable among the Spike mutations in A.2.5 are two novel INDELS, a deletion of positions 141-143 and an insertion of AGY at 215. Both indels are located in the NTD of the spike protein (Figure 4), a site of importance in the host immune response. While the receptor binding domain (RBD) of the S protein has received much of the attention due to the phenotypic impact of ACE2 receptor binding, the NTD has recently emerged as significant in its role as a site for neutralizing antibodies (37-42). Recently, three studies identified an antigenic supersite in the NTD, which includes three of five loops on the NTD at amino acid residues 14-26, 141-156, and 246-260 (39, 40, 42, 43). Notably, the deletion of residues 141-143 in A.2.5 occurs in the NTD antigenic supersite and likely interferes with neutralizing antibody binding, although future studies would need to determine the exact phenotypic effects of this deletion.

Several variants have mutations in the NTD antigenic supersite, suggesting selective pressure on this region by emerging variants to escape NTD-specific neutralizing antibodies (39, 40, 42, 43). Additional mutations causing increased transmissibility are likely key factors in why variants, such as Alpha (B.1.1.7) and Delta (B.1.671.2), have significant case counts around the world and soon became the dominant variant in geographical areas, such as the U.S., India, and the U.K. shortly after they emerged (1, 11, 44, 45). While A.2.5 does not appear to demonstrate increased transmissibility based on the frequency of the lineage in the U.S. and globally, A.2.5 and its emerging sublineages should continue to be monitored as novel mutations could arise to increase transmissibility.

A.2.5 experienced rapid global expansion early during its emergence, however cases in Florida have been generally low to-date, especially in comparison to other variants (e.g., B.1.1.7, P.1, and B.1.617.2). Lineages with concerning or novel mutations in the NTD or RBD of spike must be further investigated and monitored to determine their phenotypic effects on transmissibility or therapeutics. Case counts and lineage proportions are only one factor when considering the effect of a particular variant on distinct populations within a community. As vaccination rates increase, restrictions continue to lift, and domestic and international travel increase, transmission dynamics will alter, and selective pressures may give rise to additional emerging variants or increase transmission frequencies of currently circulating variants. Continued baseline and targeted surveillance, in addition to contact tracing, are imperative to implement a strong public health response to reduce or eliminate transmission of SC2 concerning variants.

## Data Availability

All data used in the manuscript are available online. Links and accessions are in the manuscript.

## Acknowledgements

This analysis was funded in part through the Centers for Disease Control and Prevention’s Office of Advanced Molecular Detection’s support of the Epidemiology and Laboratory Capacity for Prevention and Control of Emerging Infectious Diseases Cooperative Agreement. We gratefully acknowledge the authors from the originating and submitting laboratories that provided sequences to GISAID’s EpiCoV database that were used in this study as background global samples (Supplemental Table 1).

---

**Supplemental Table 1.**
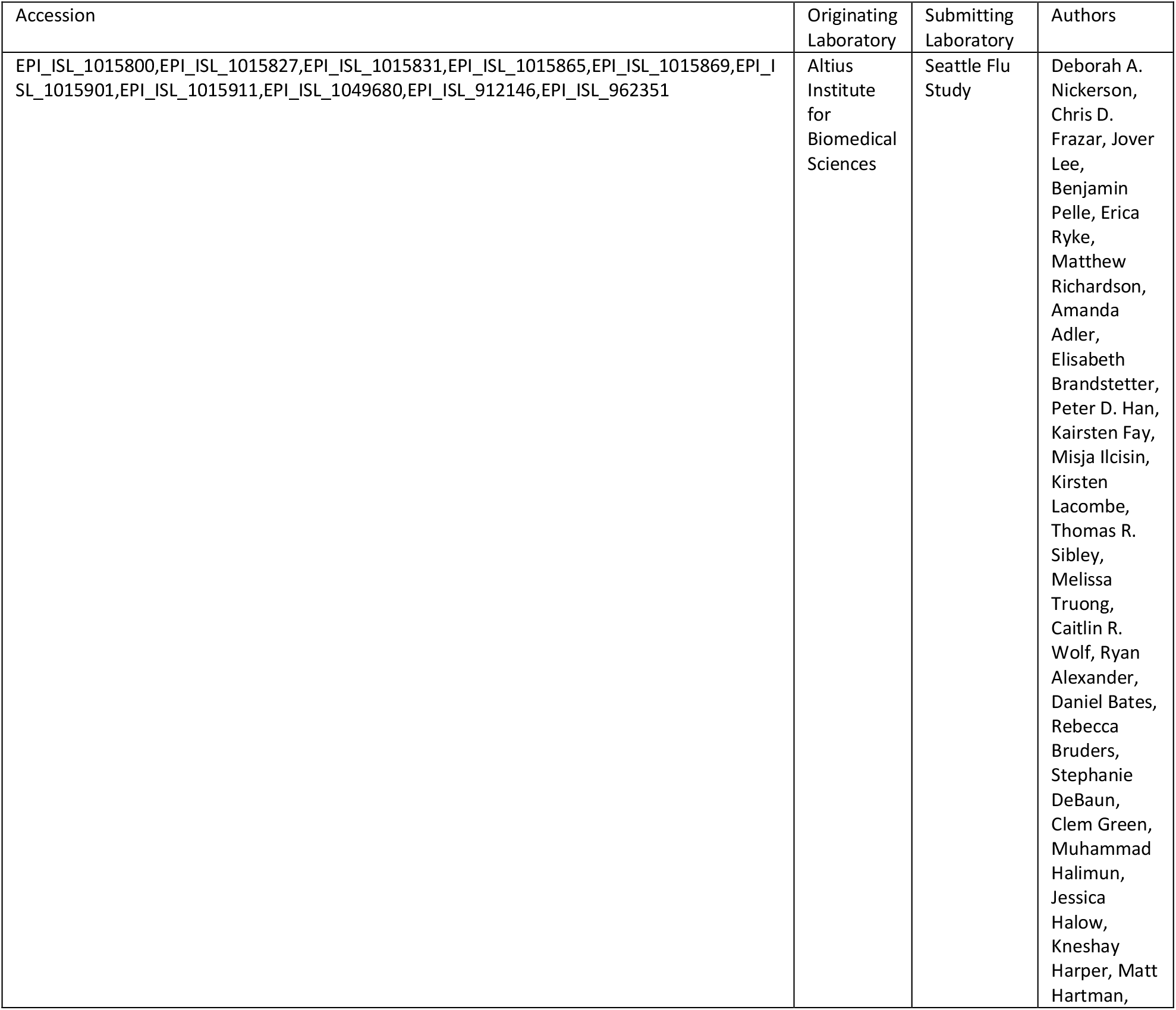
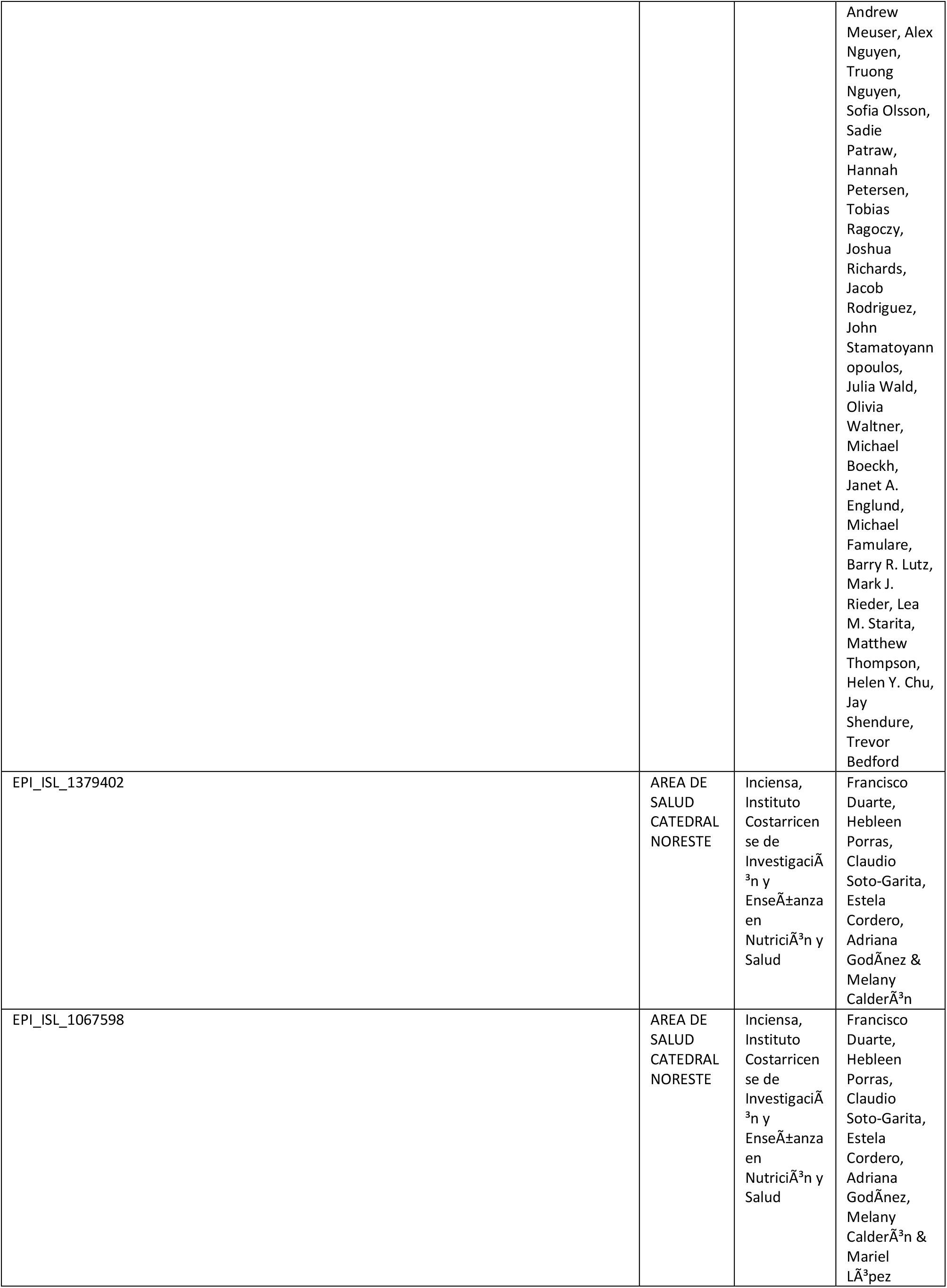
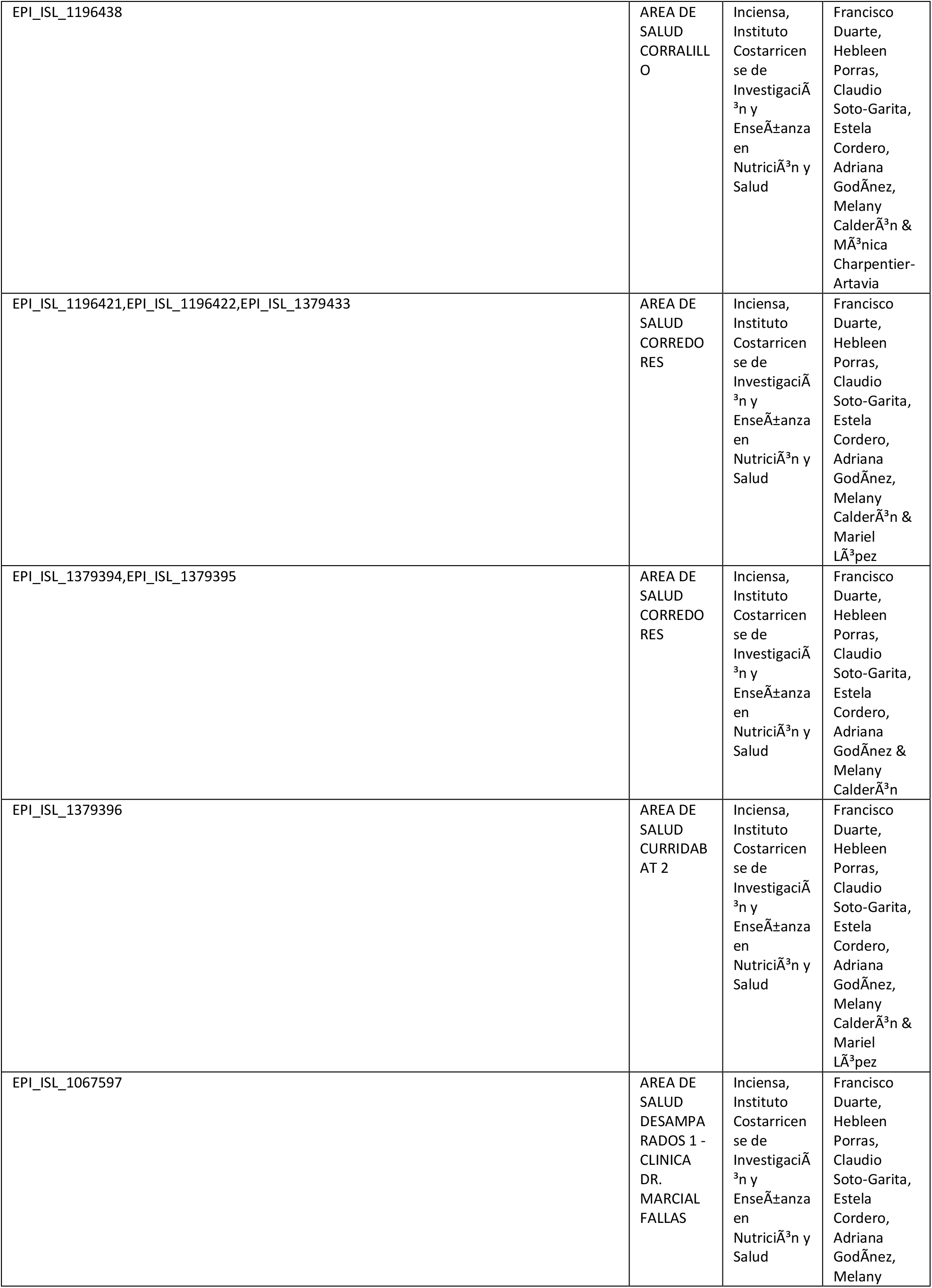
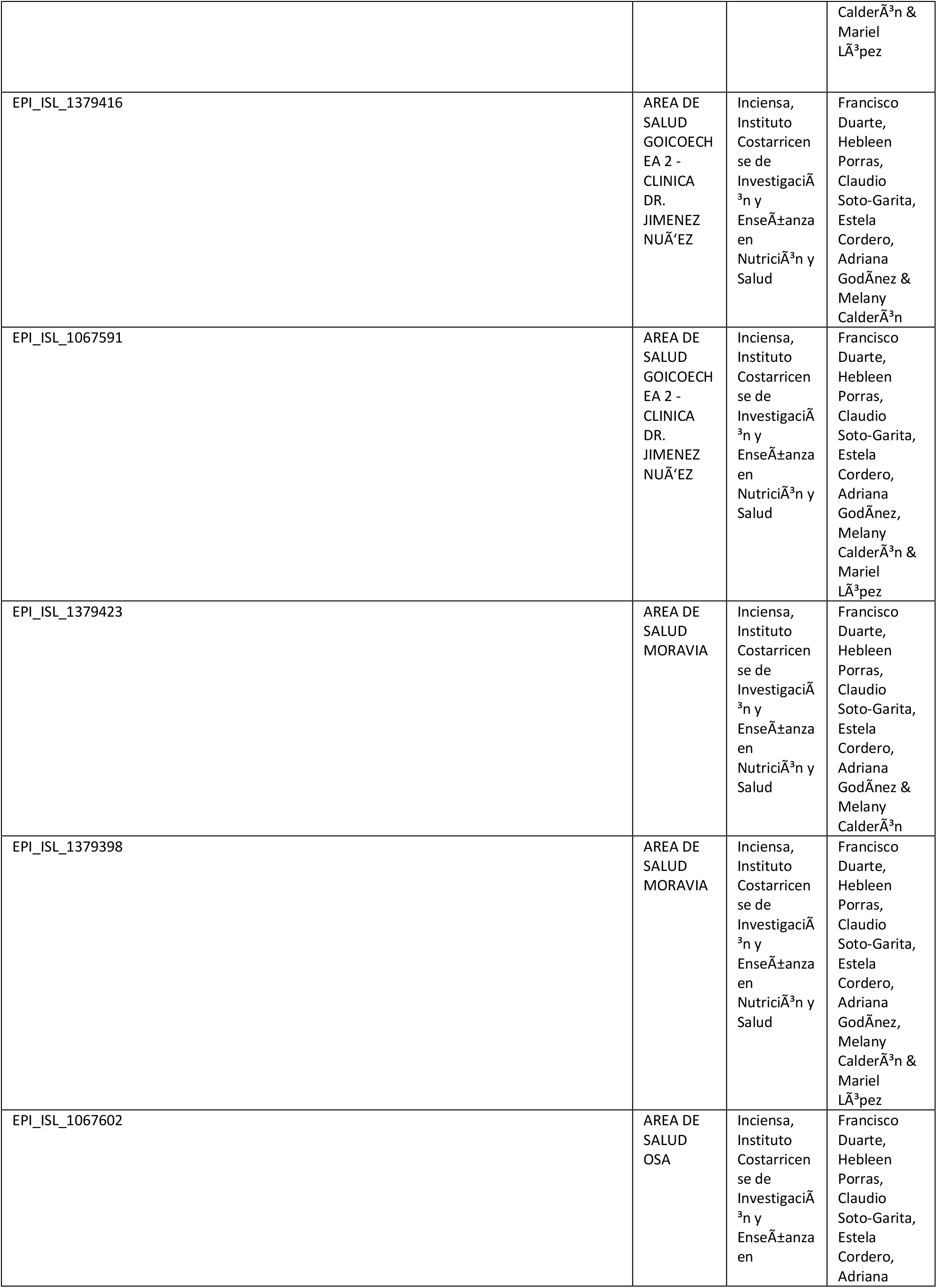
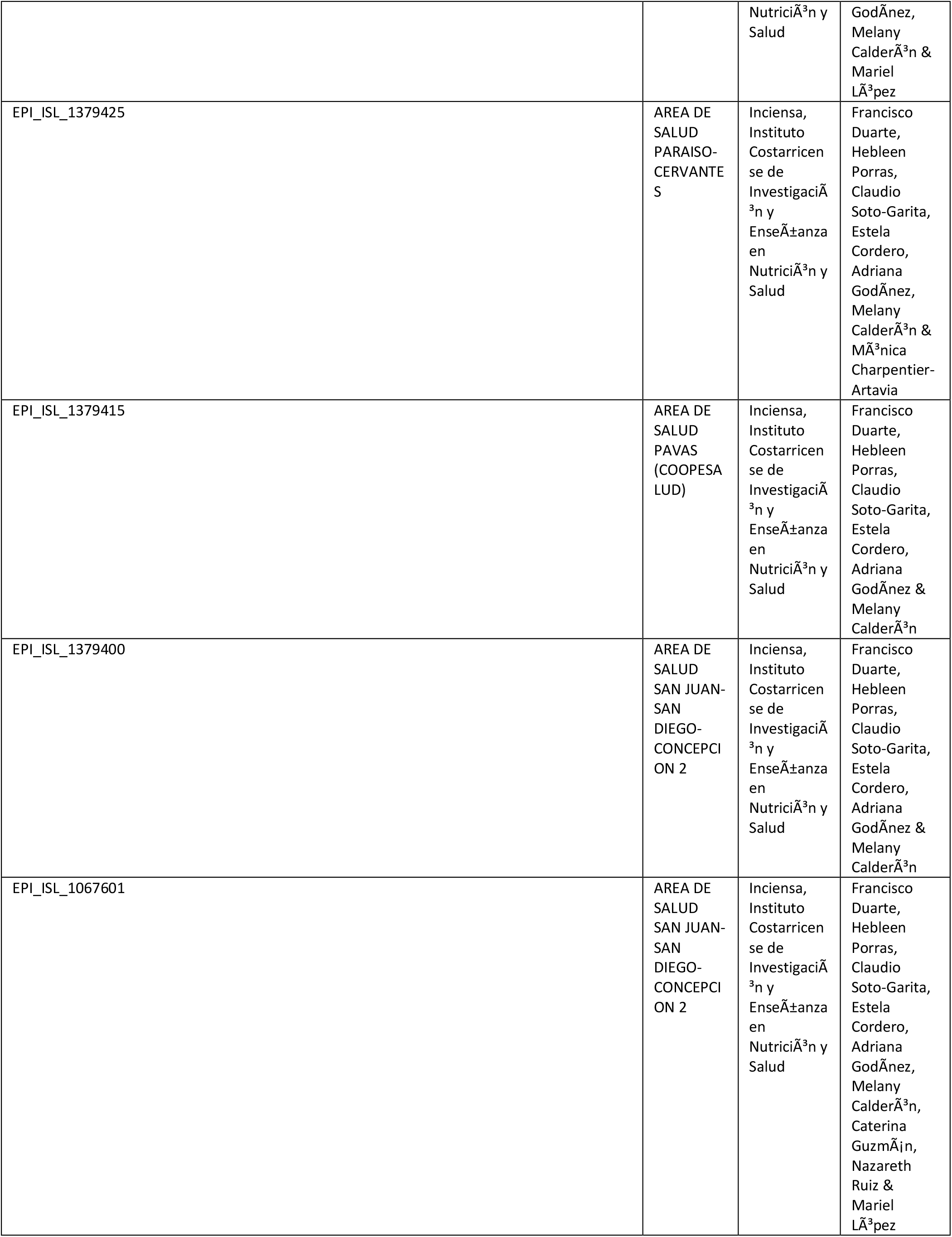
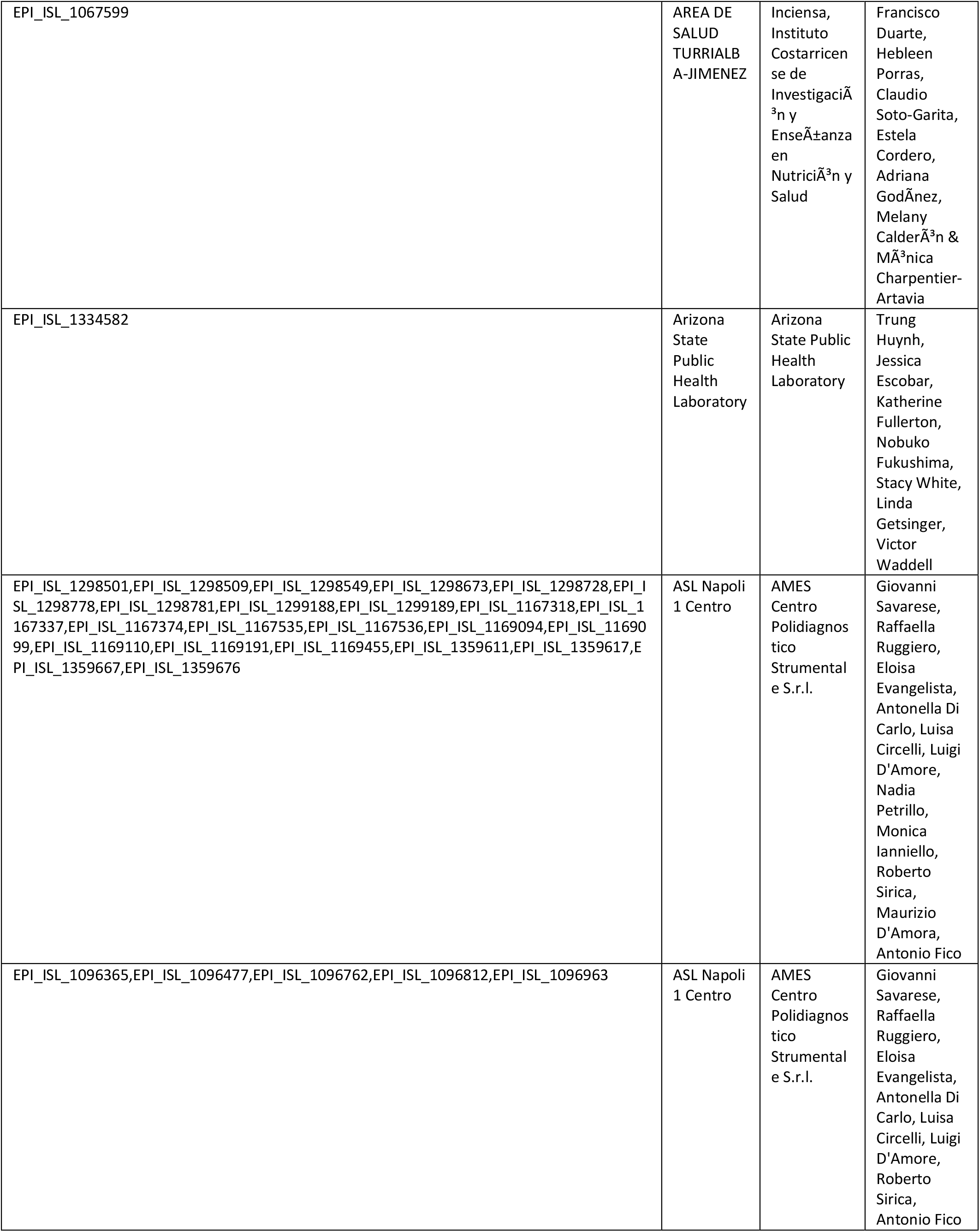
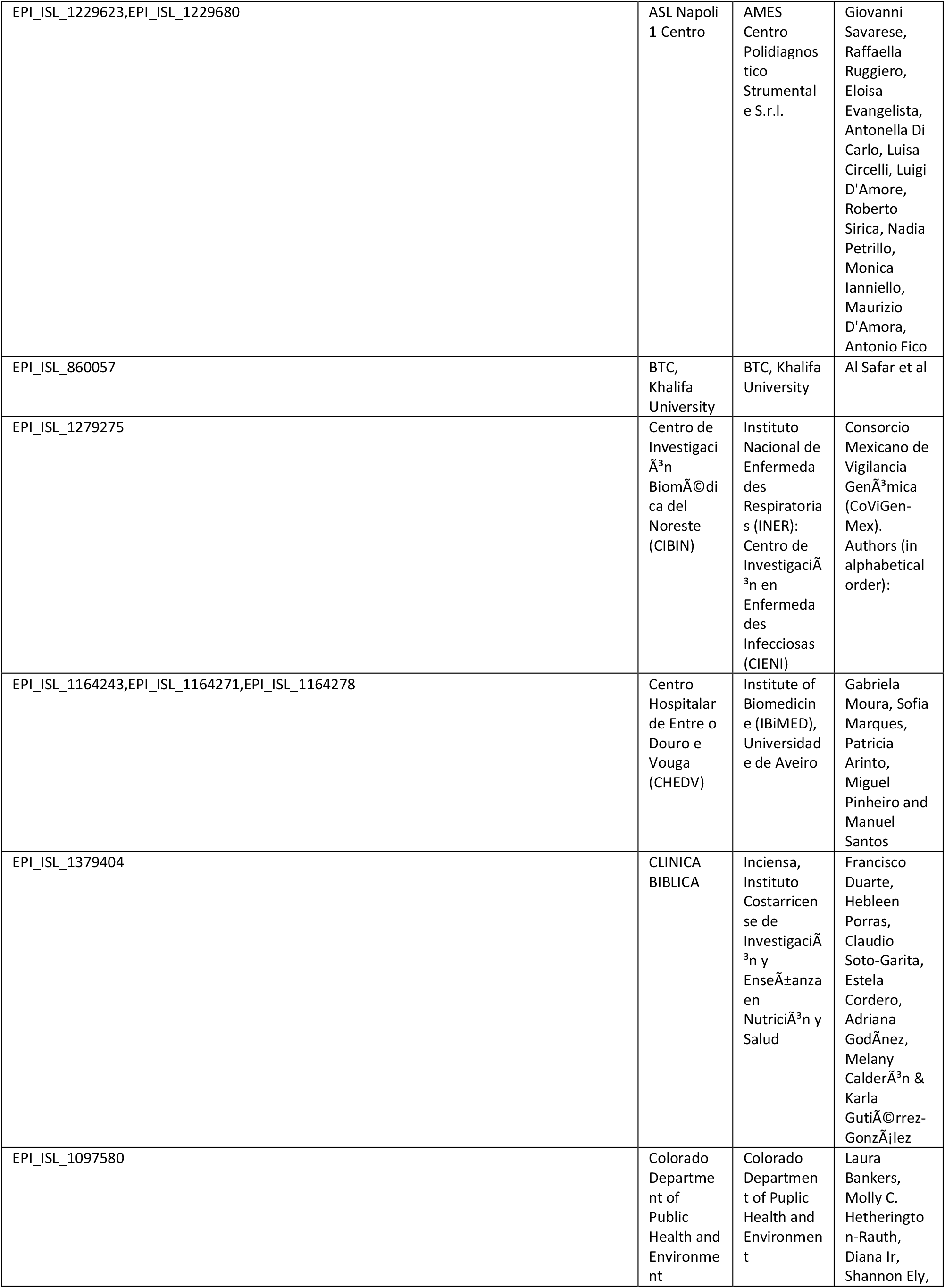
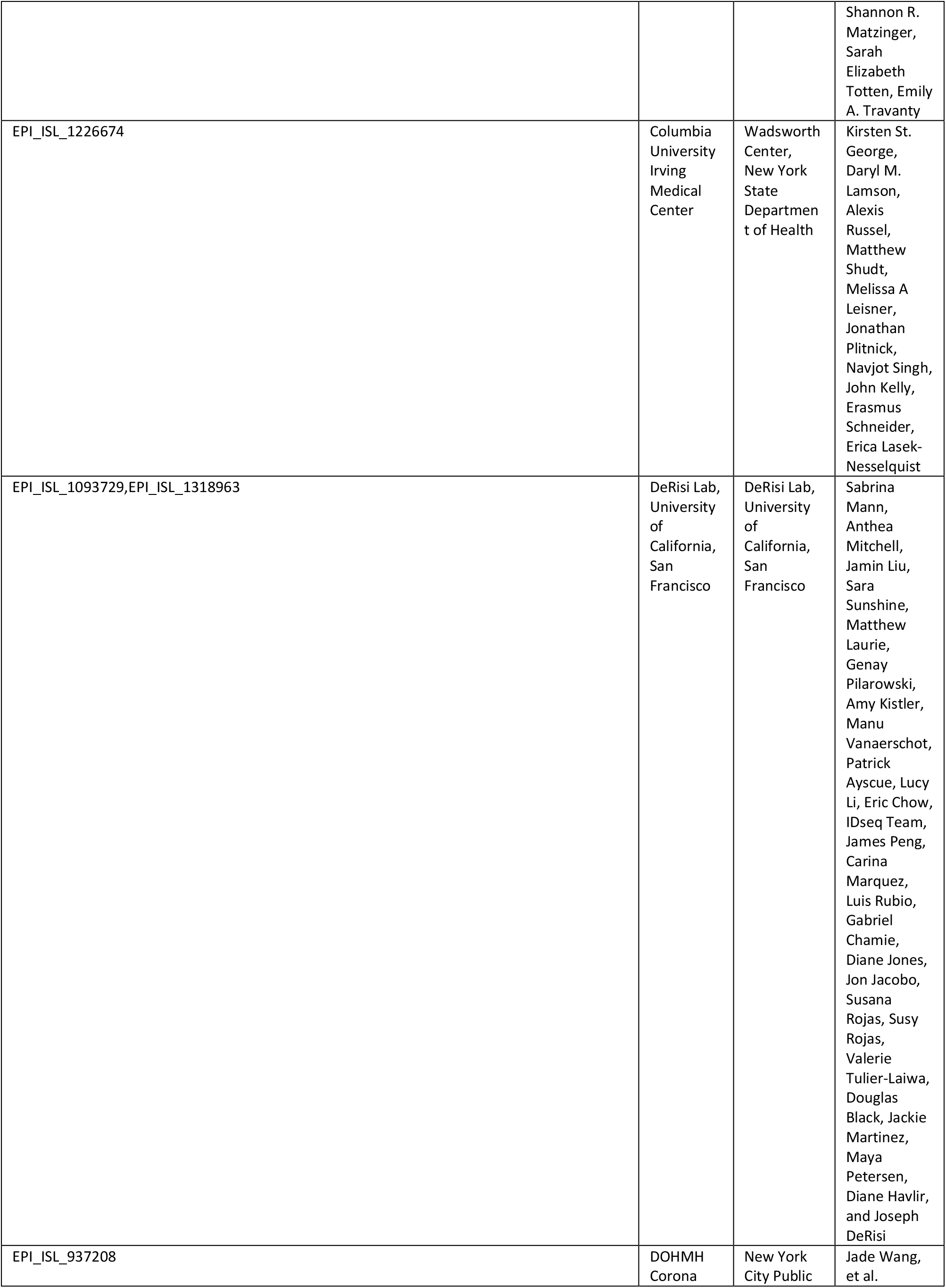
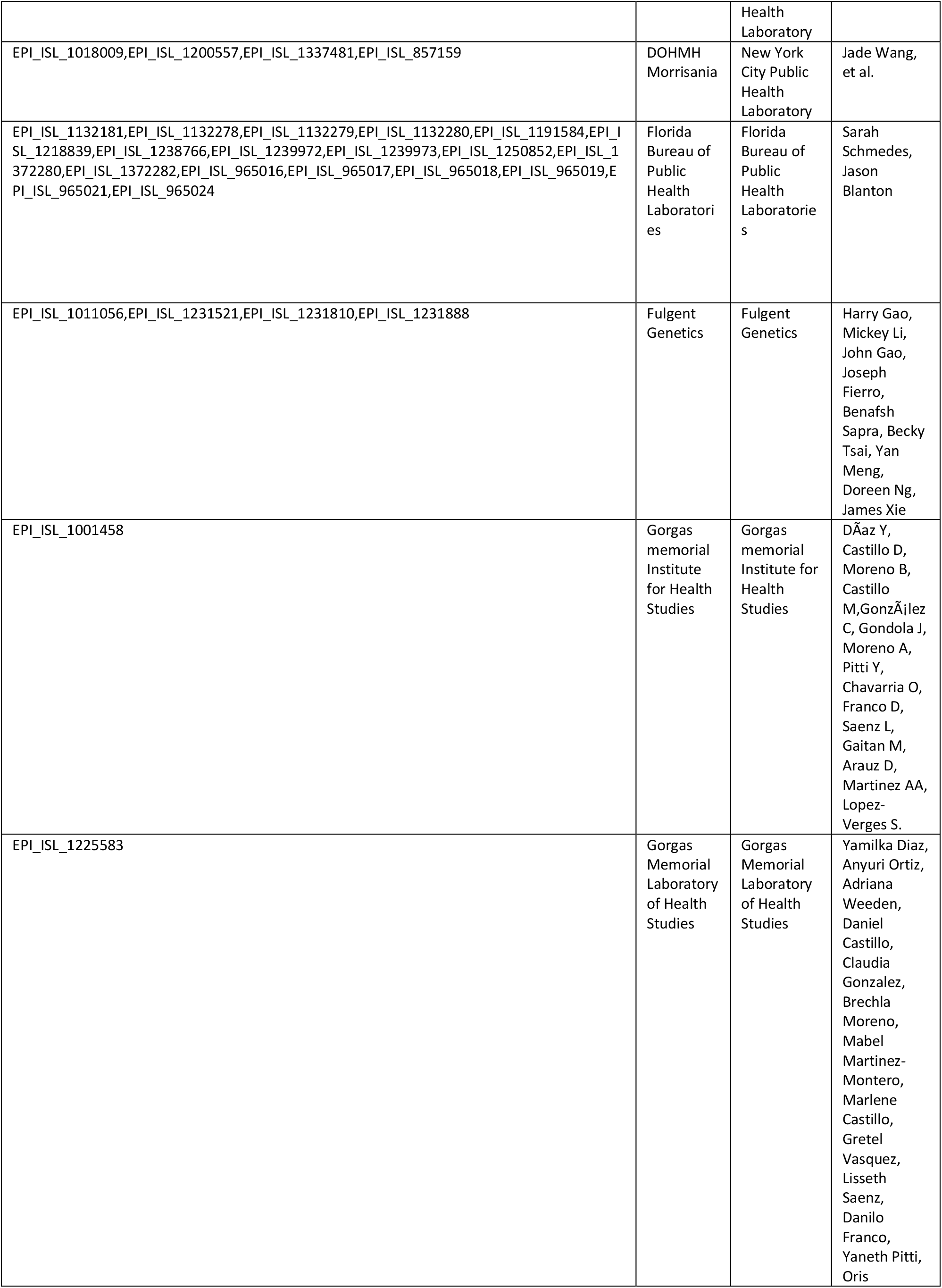
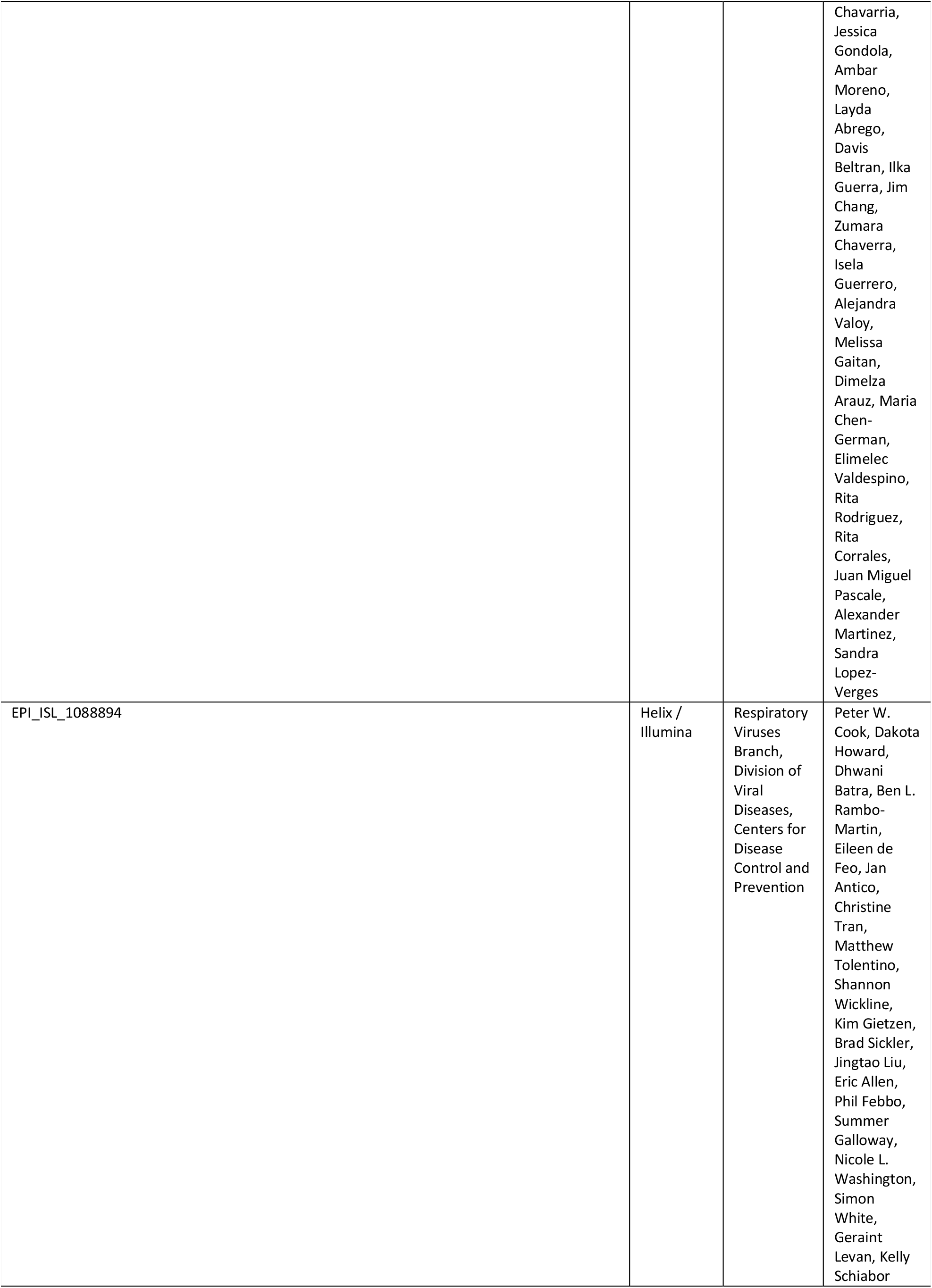
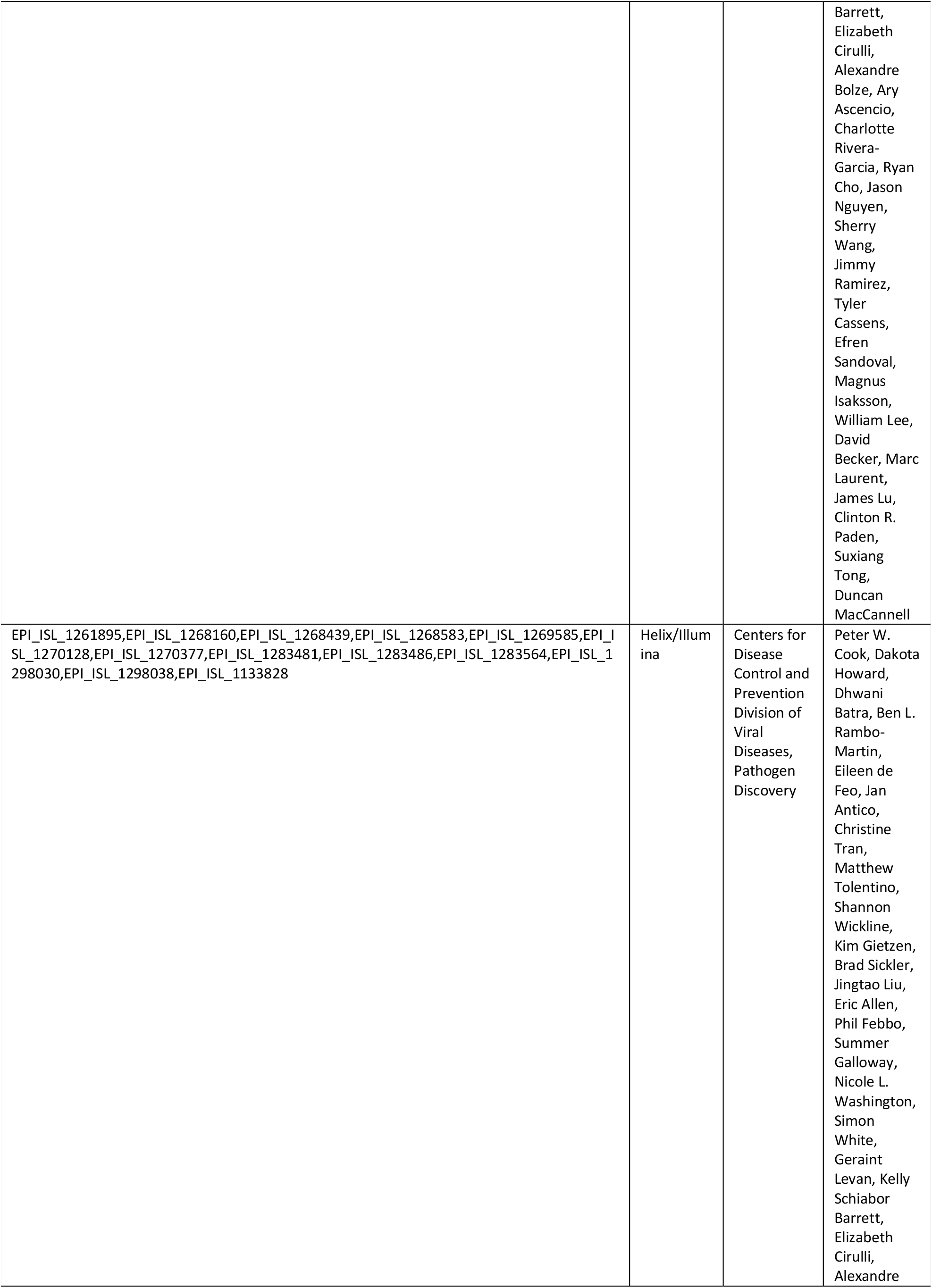
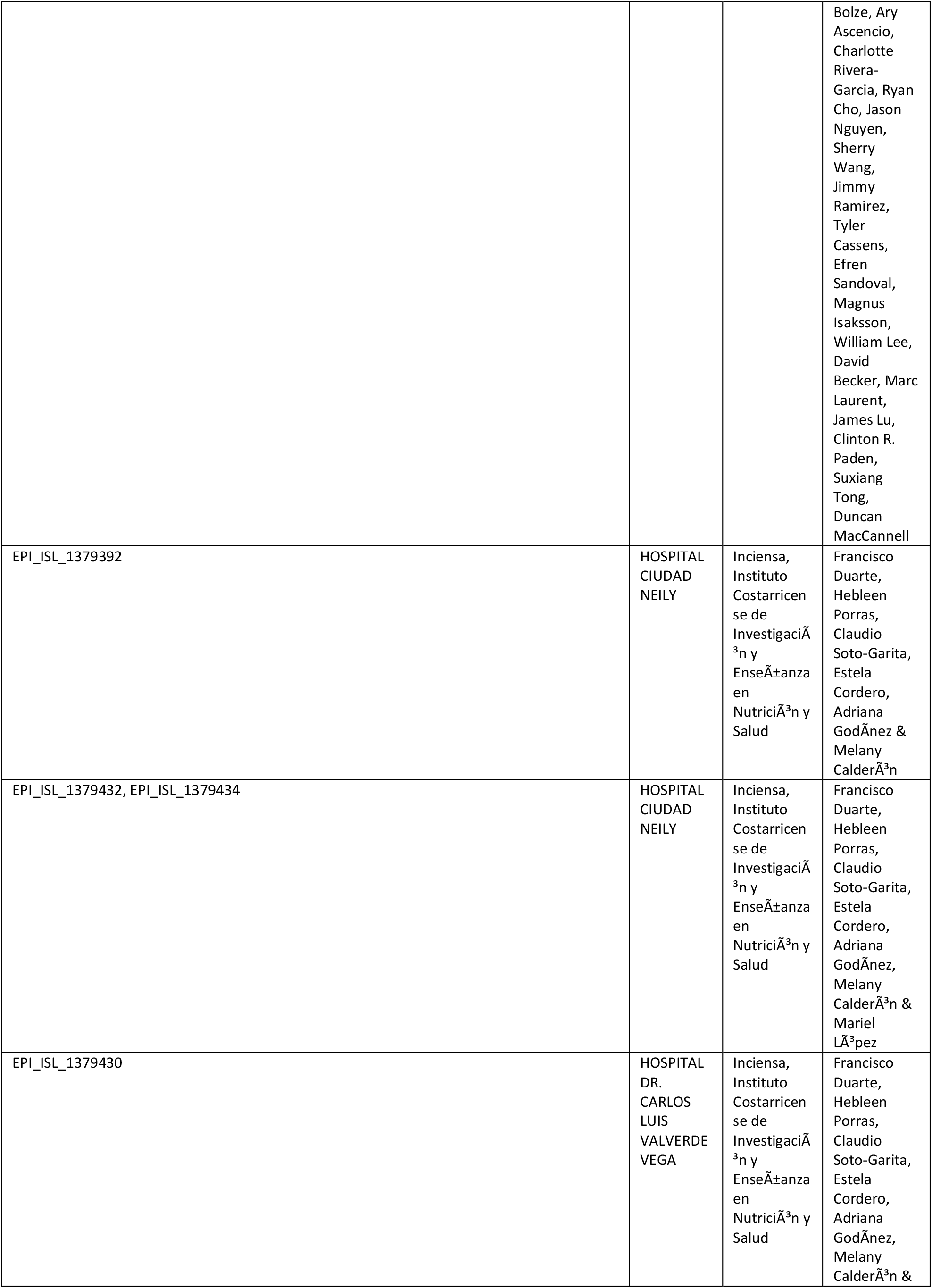
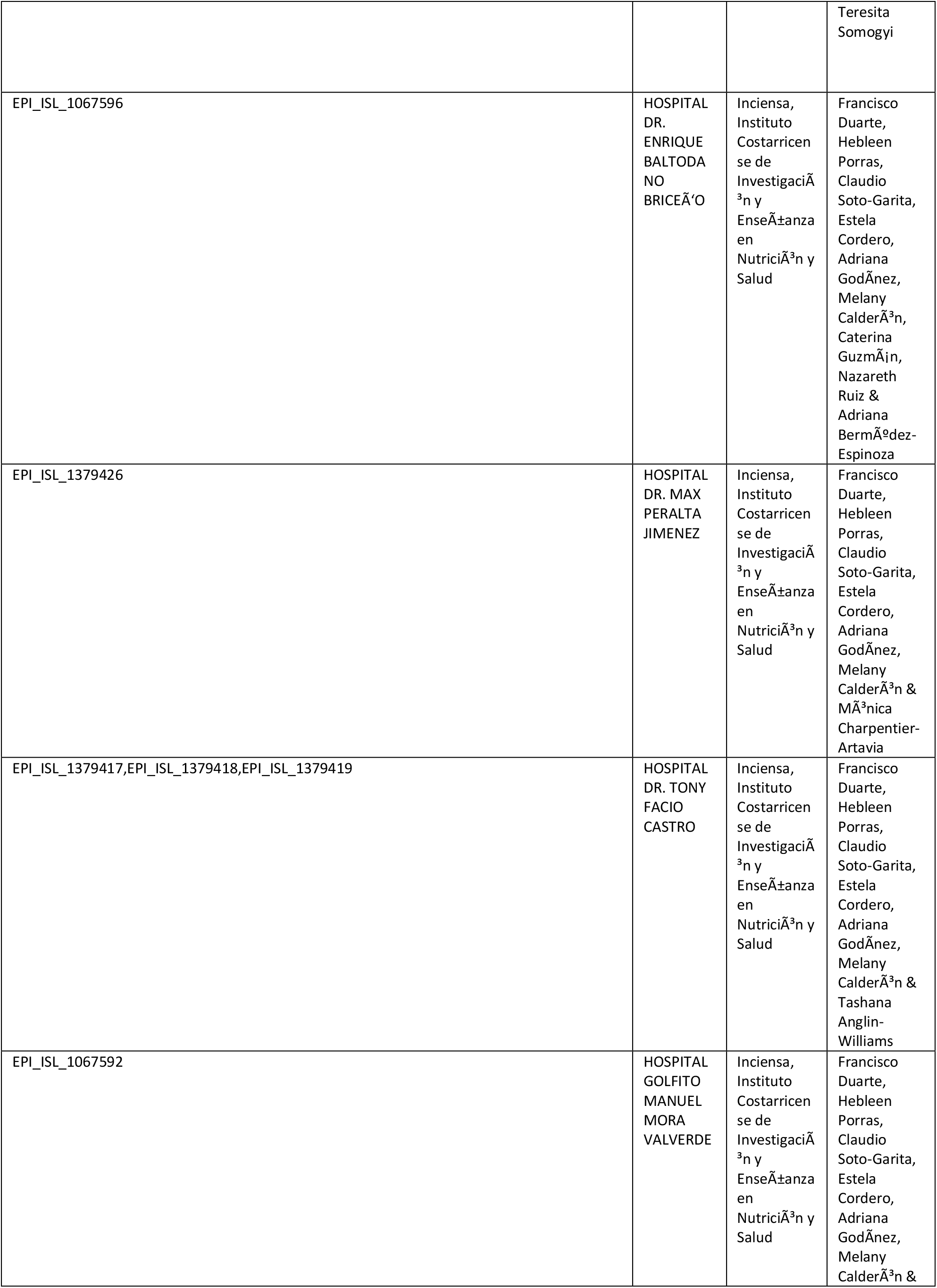
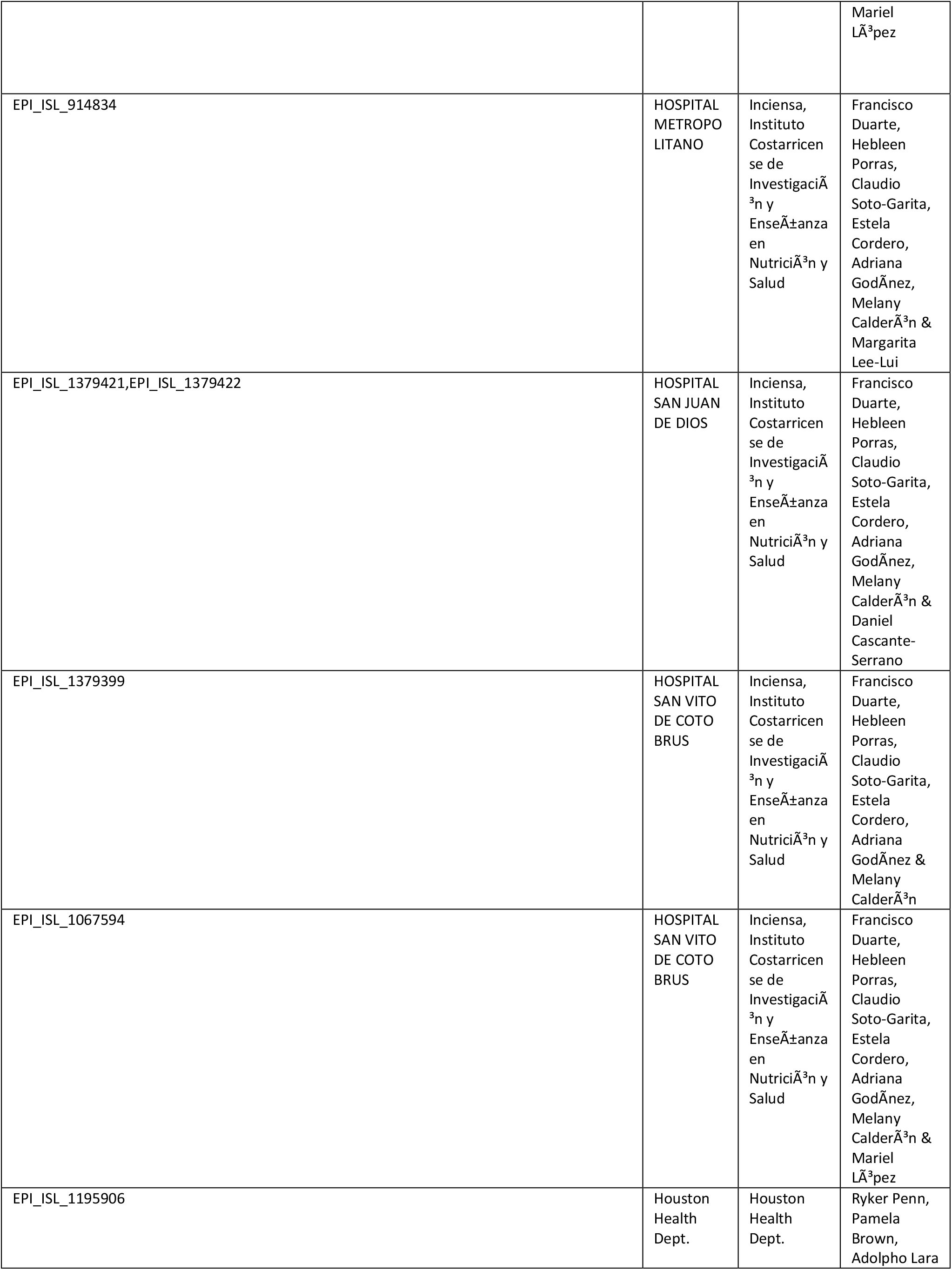
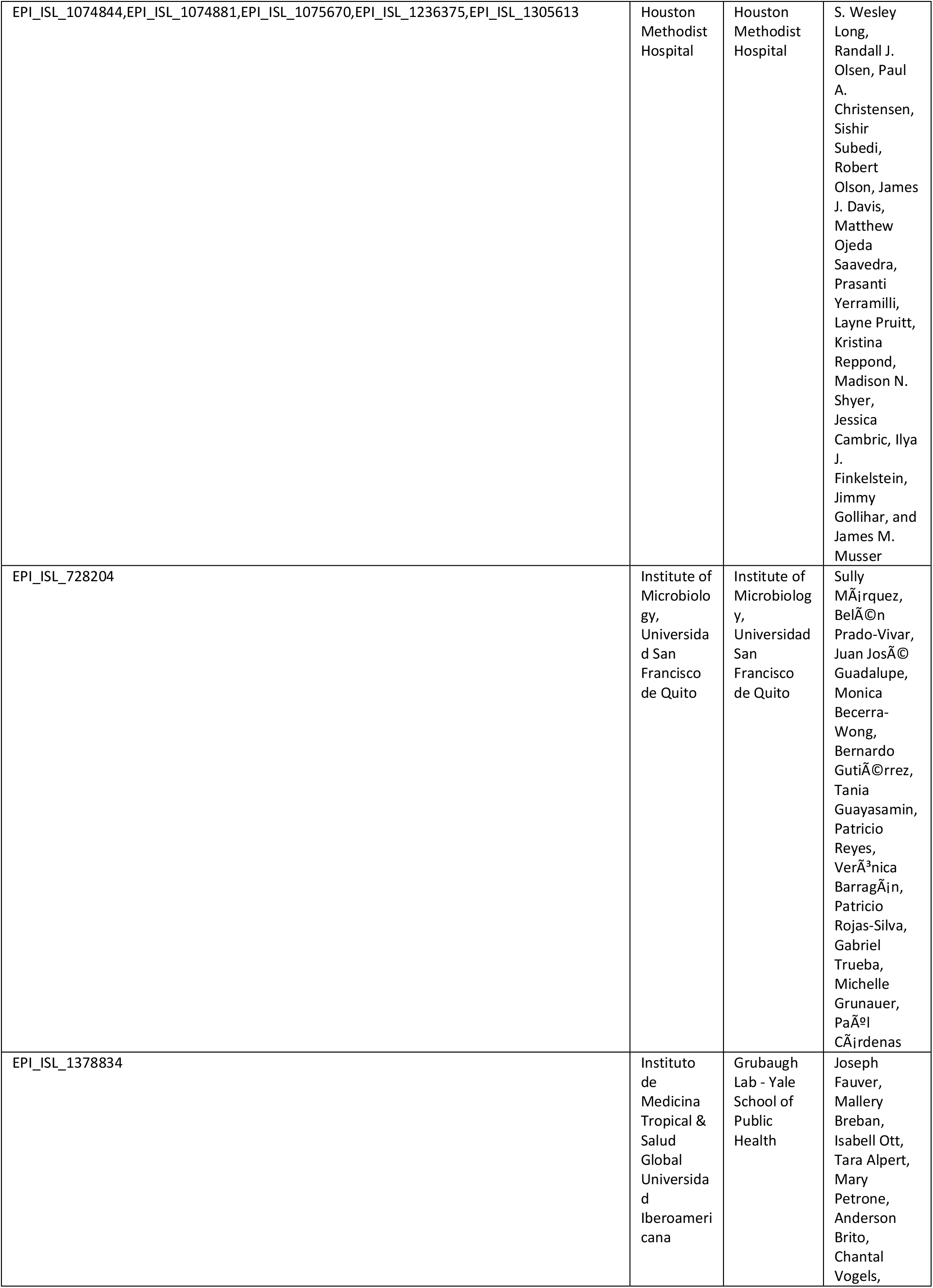
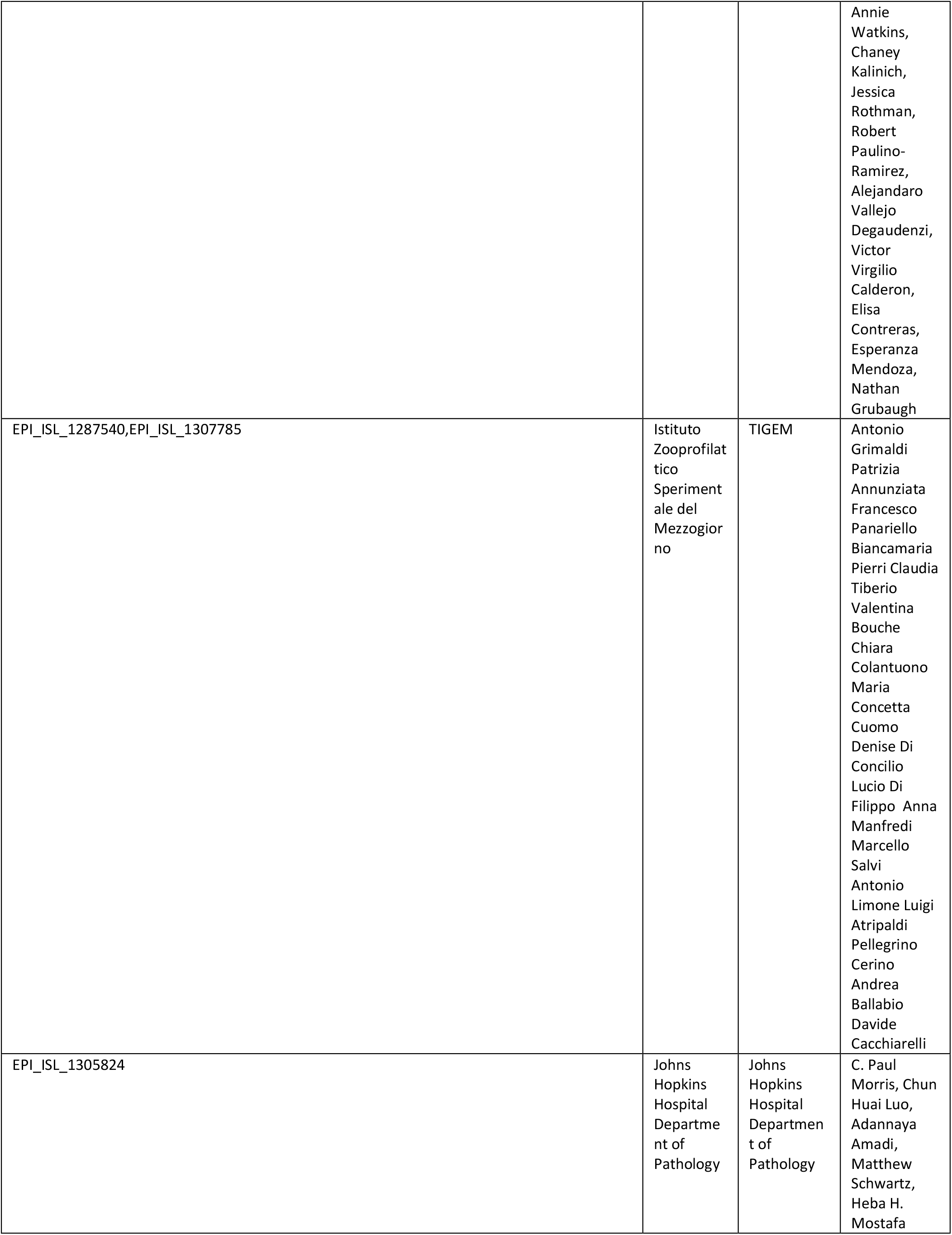
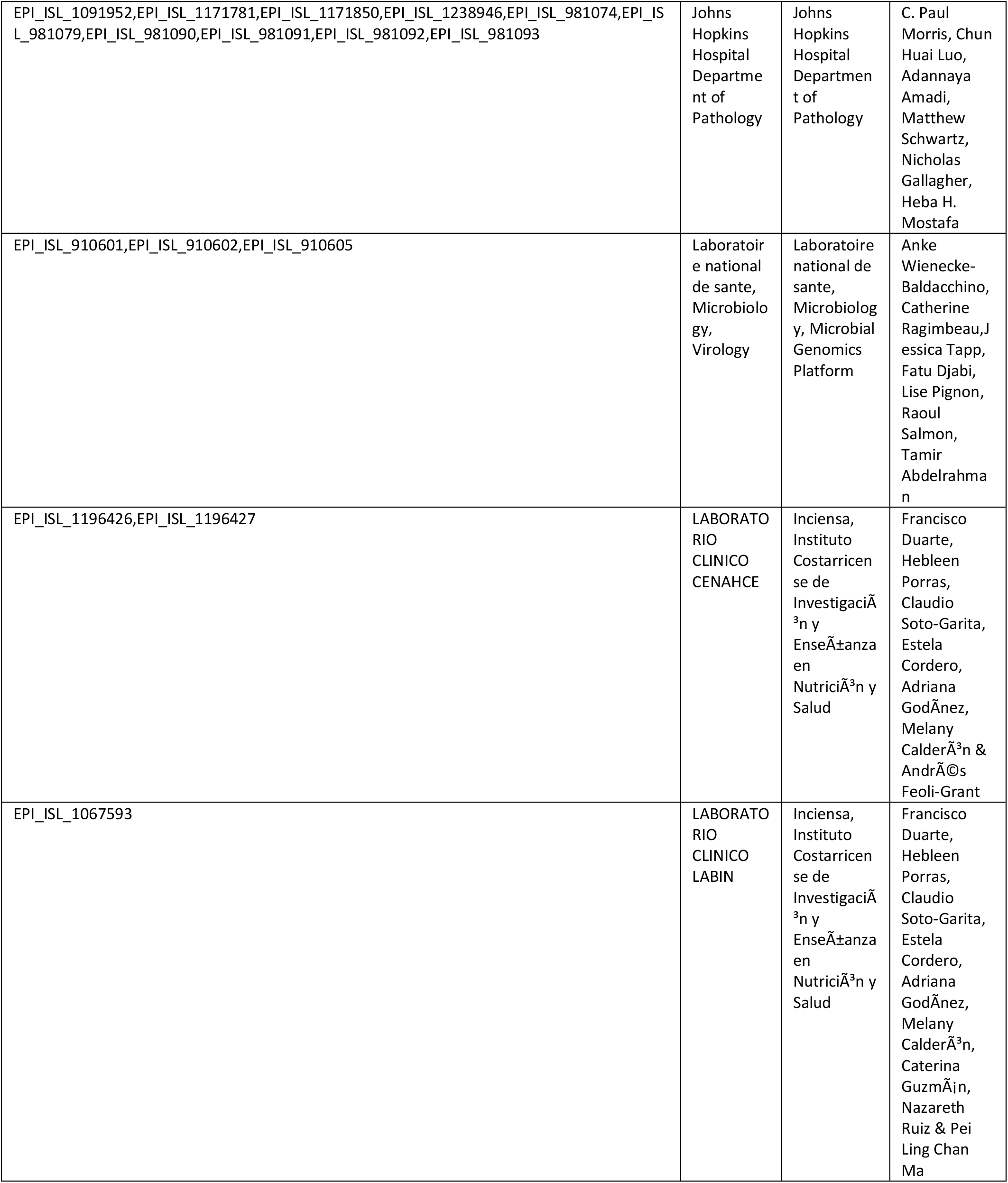
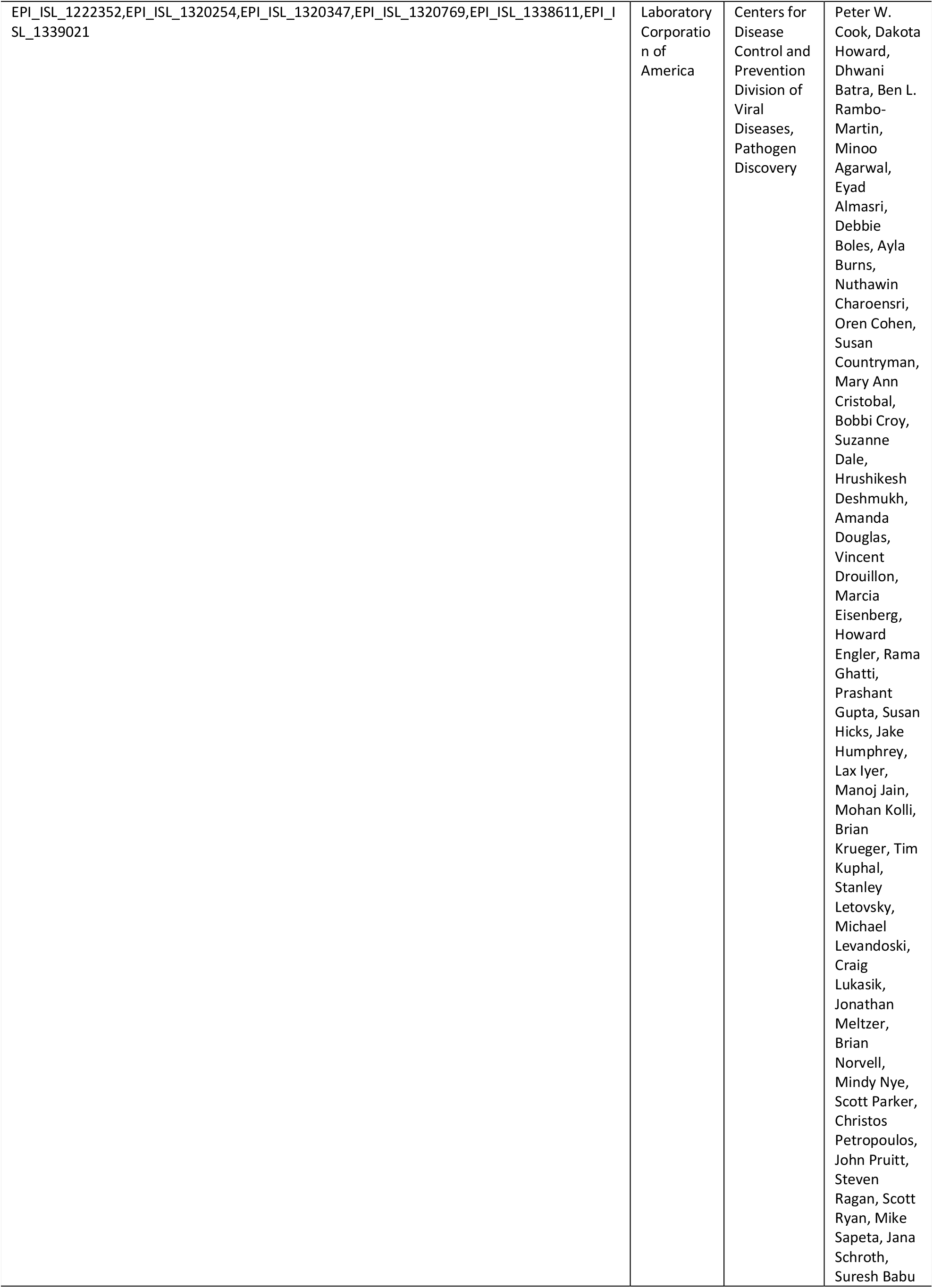
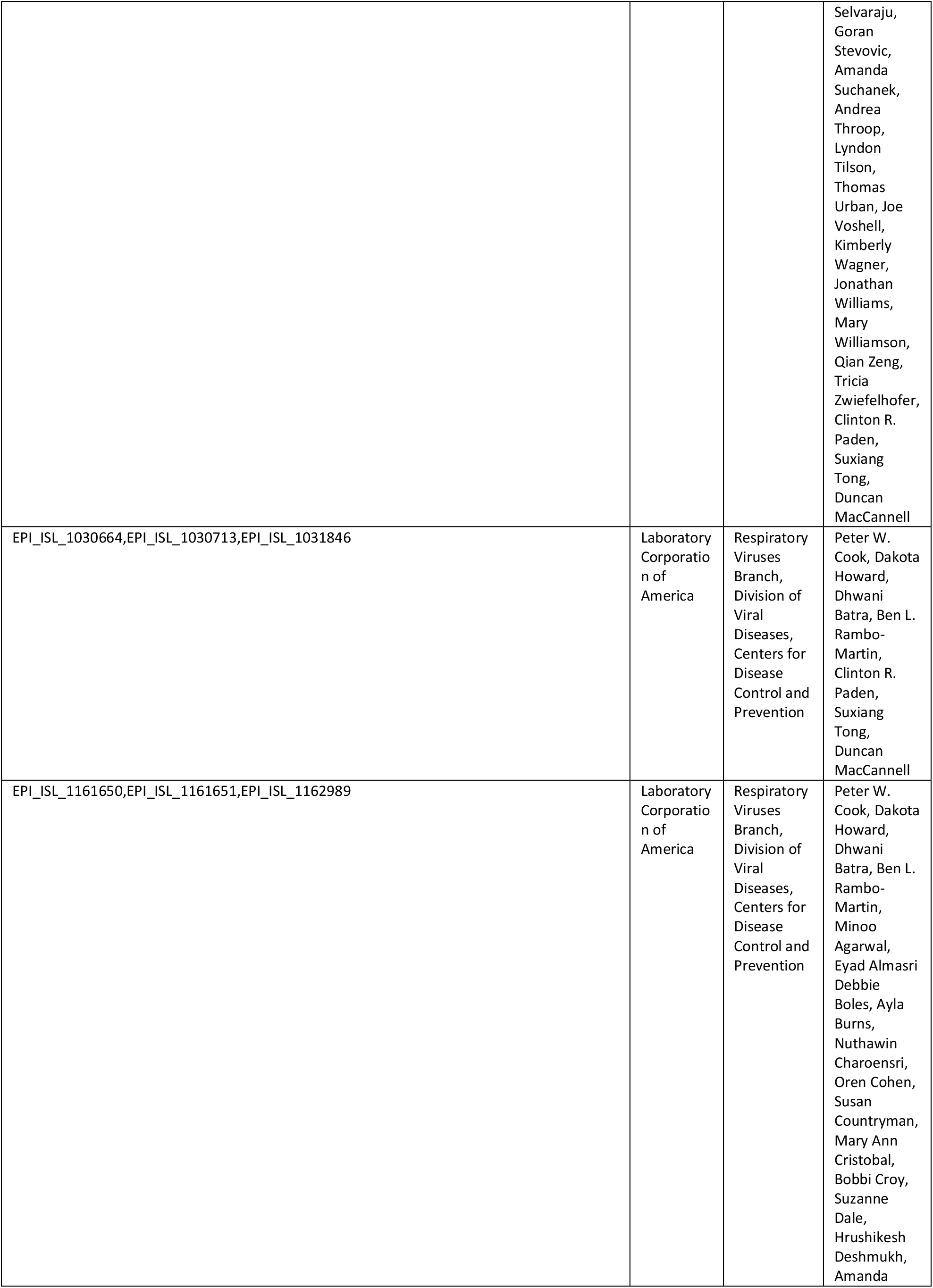
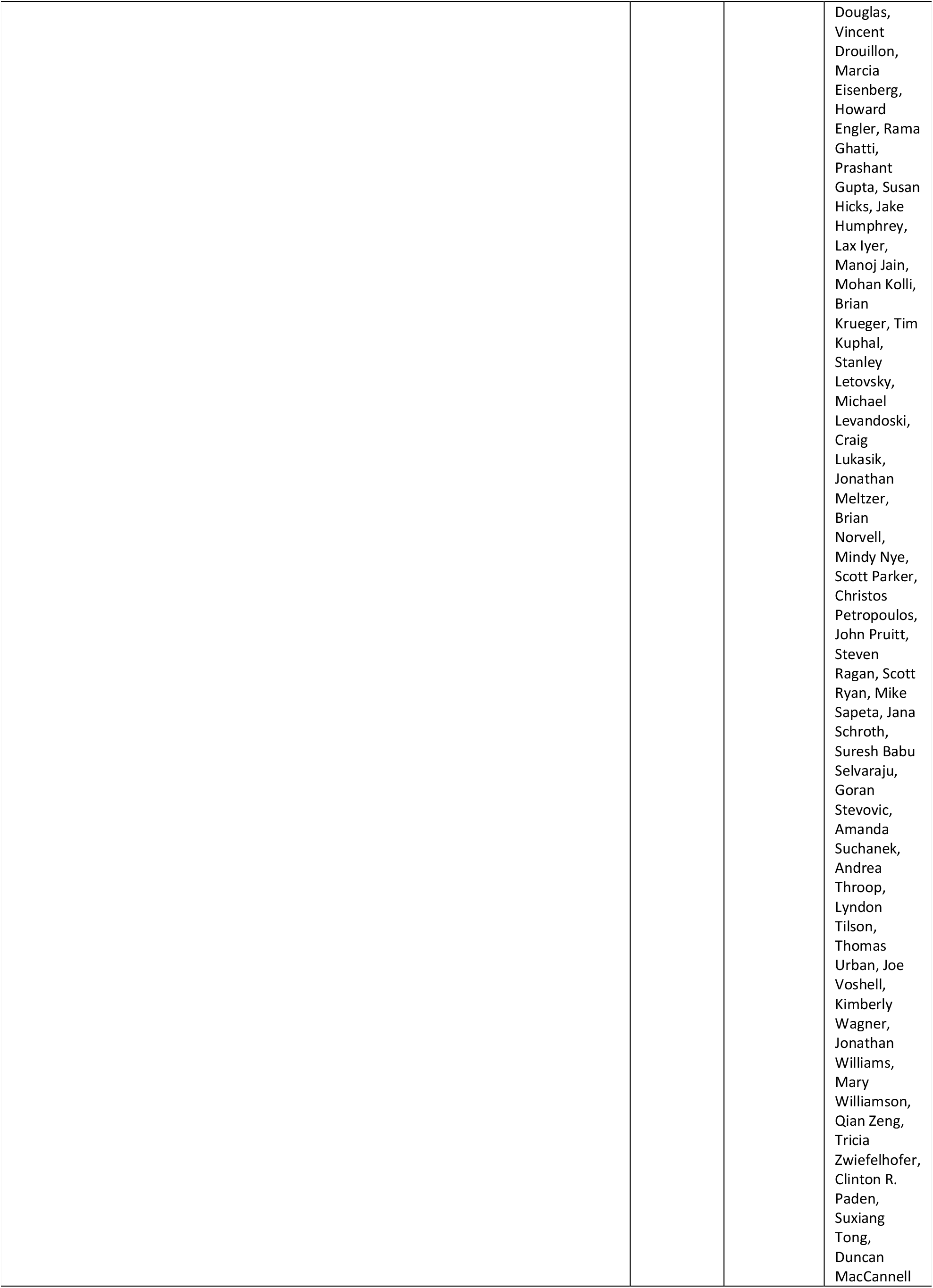
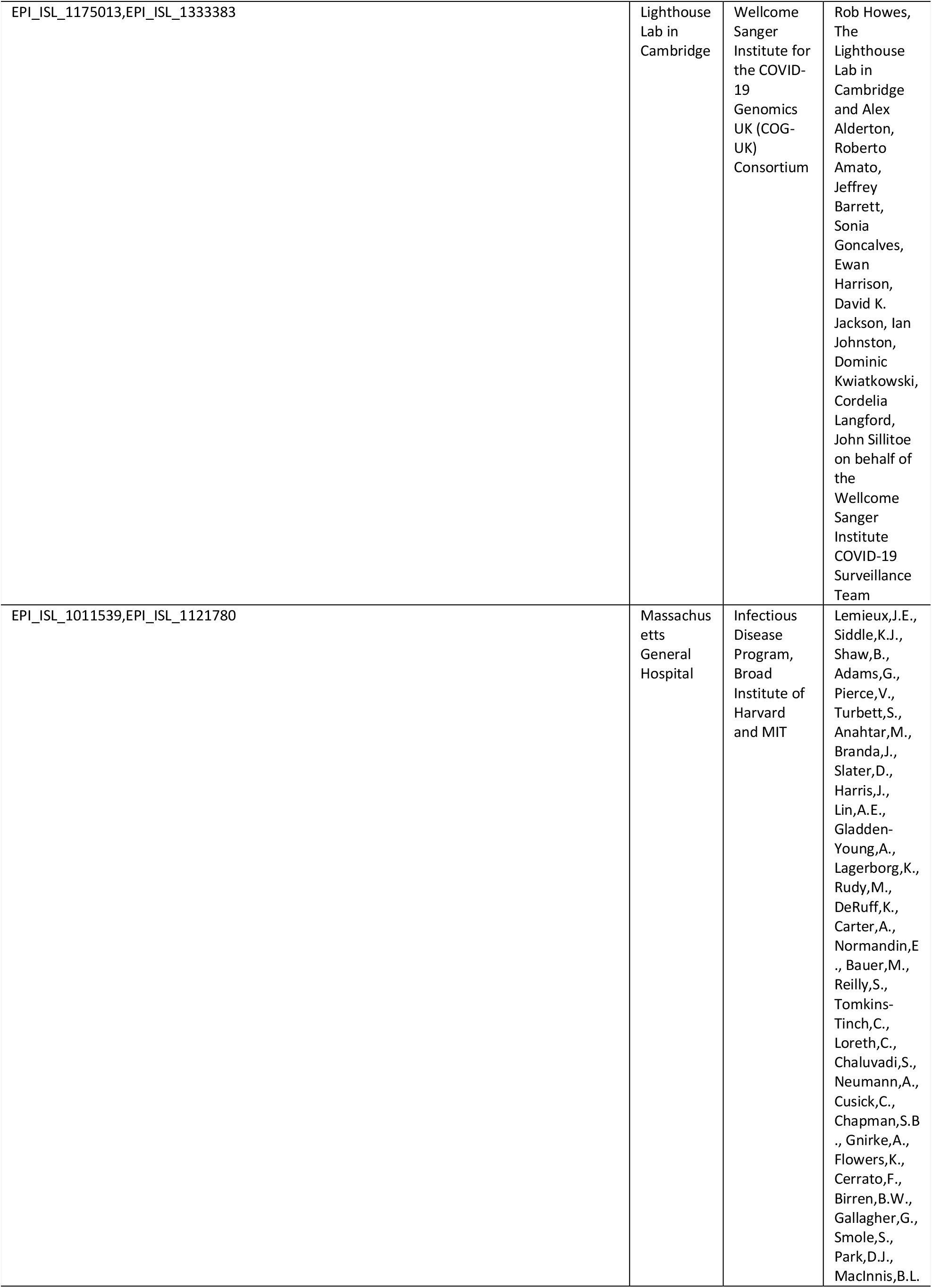
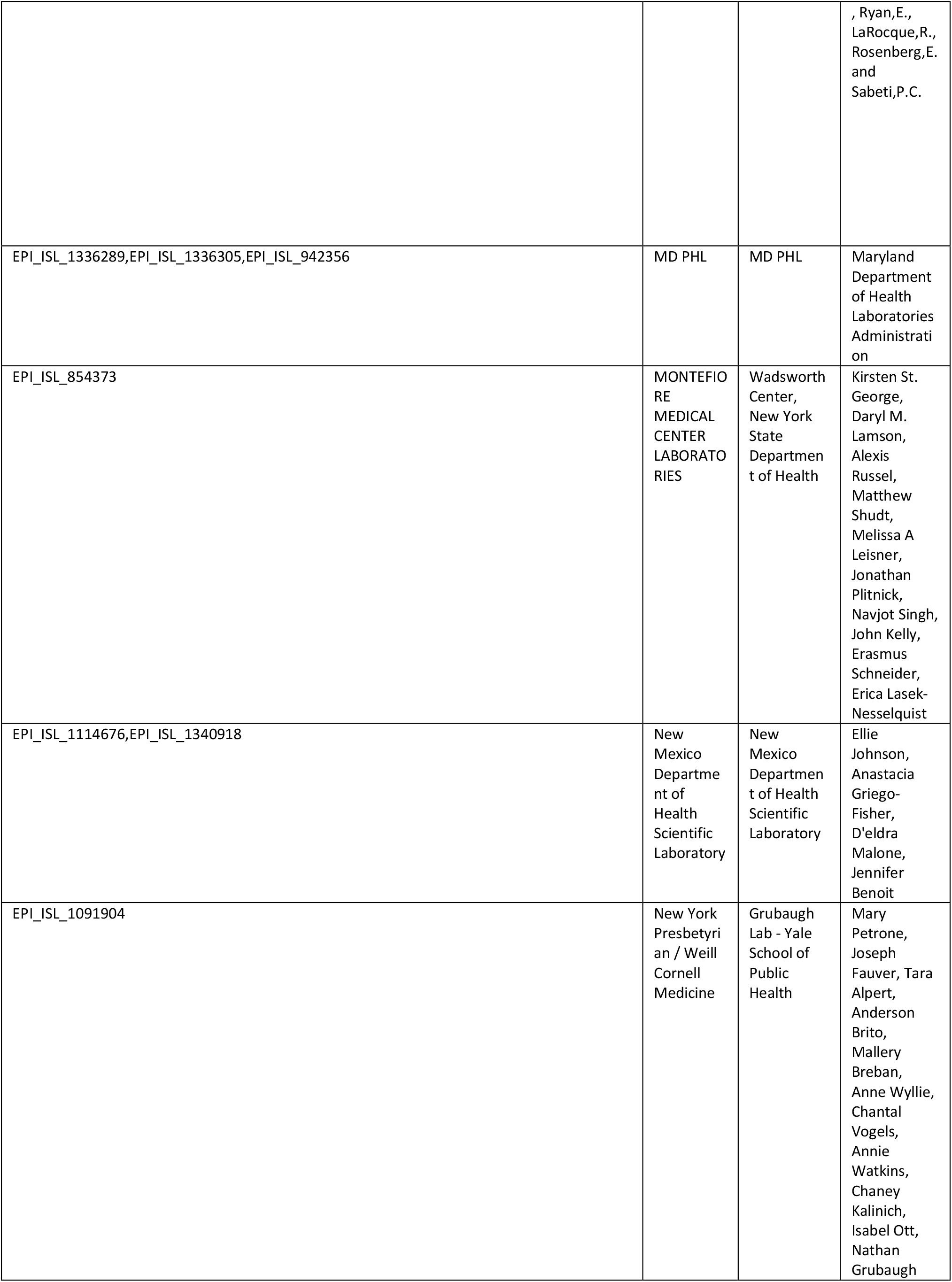
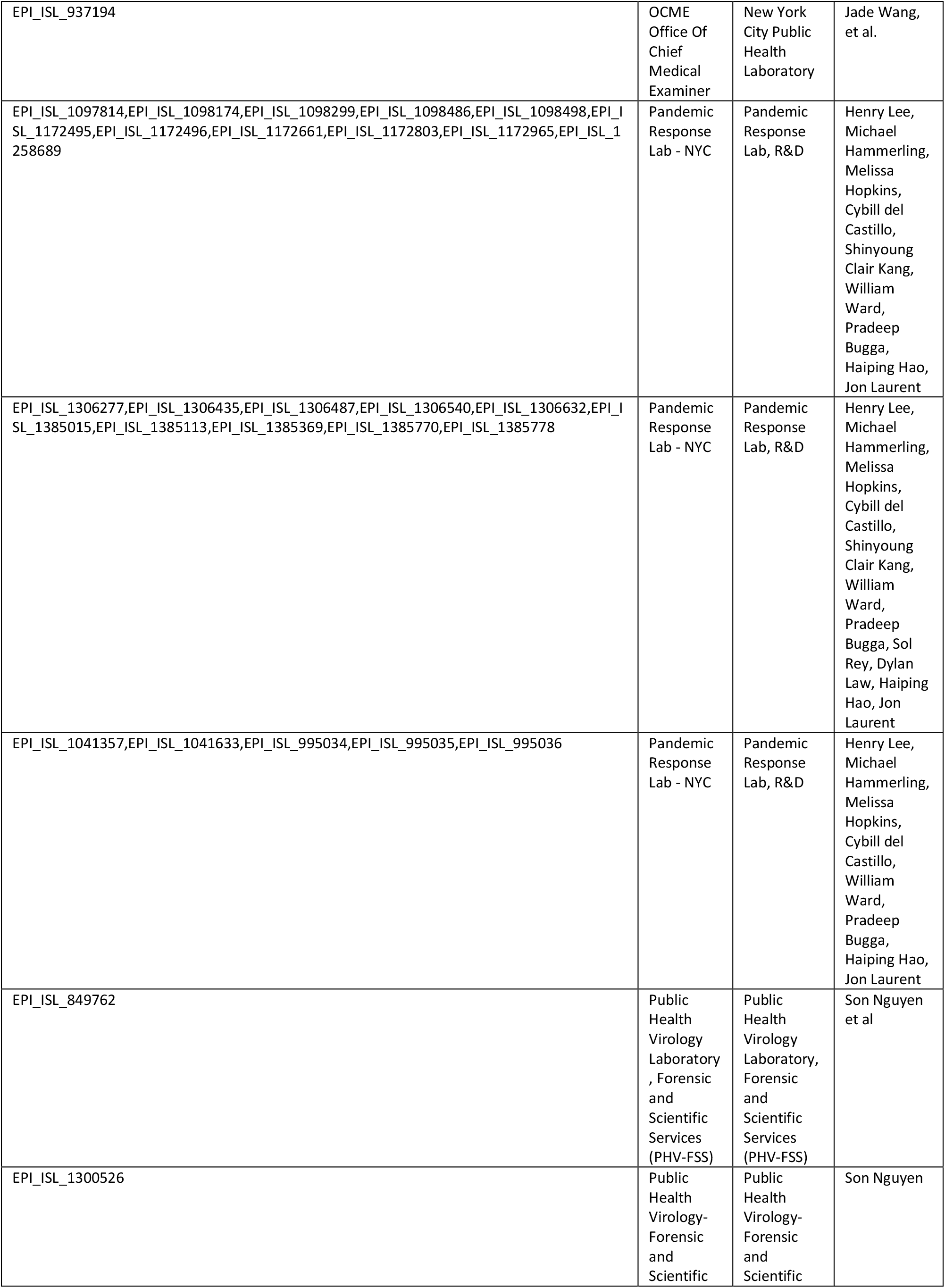
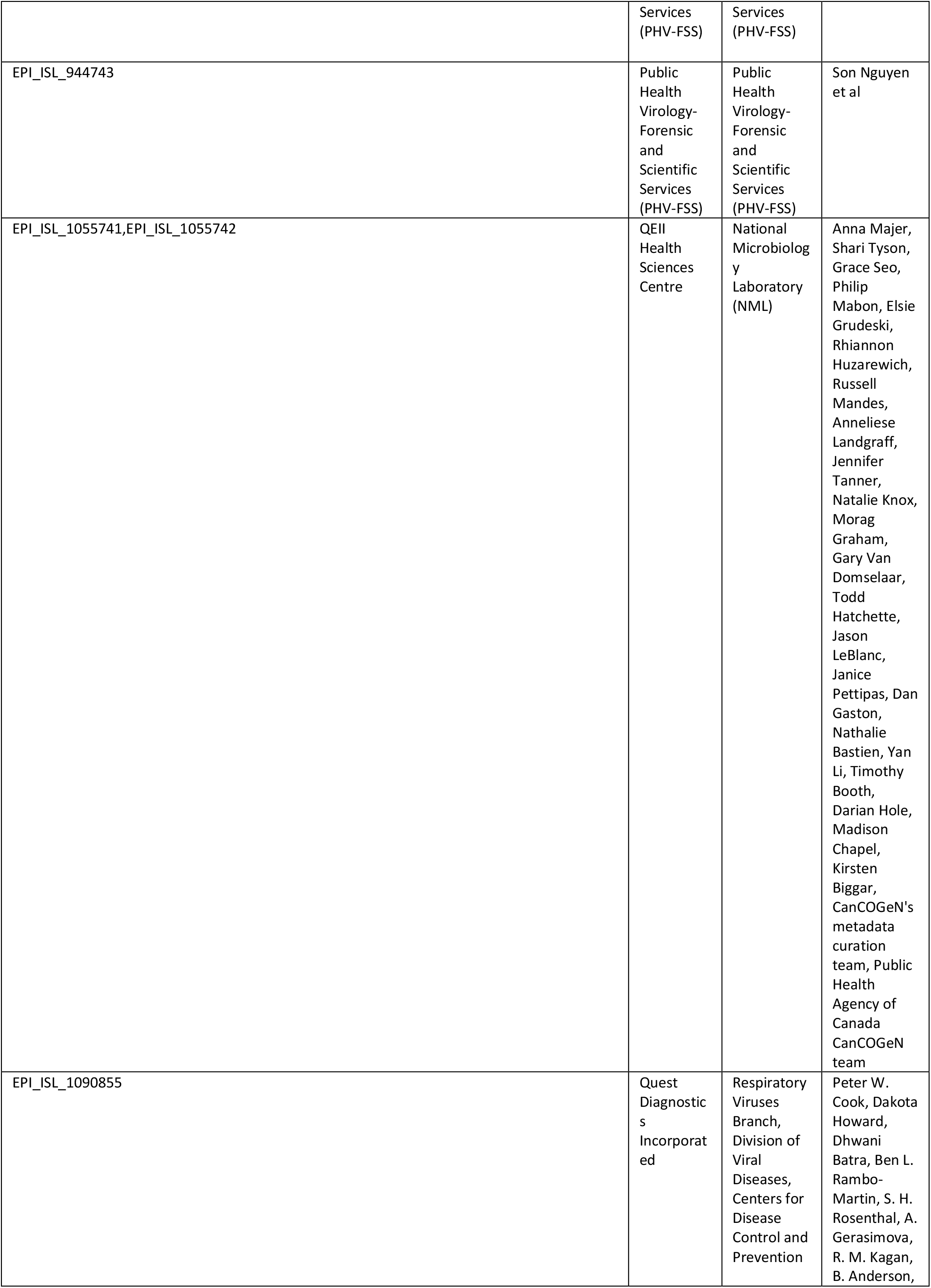
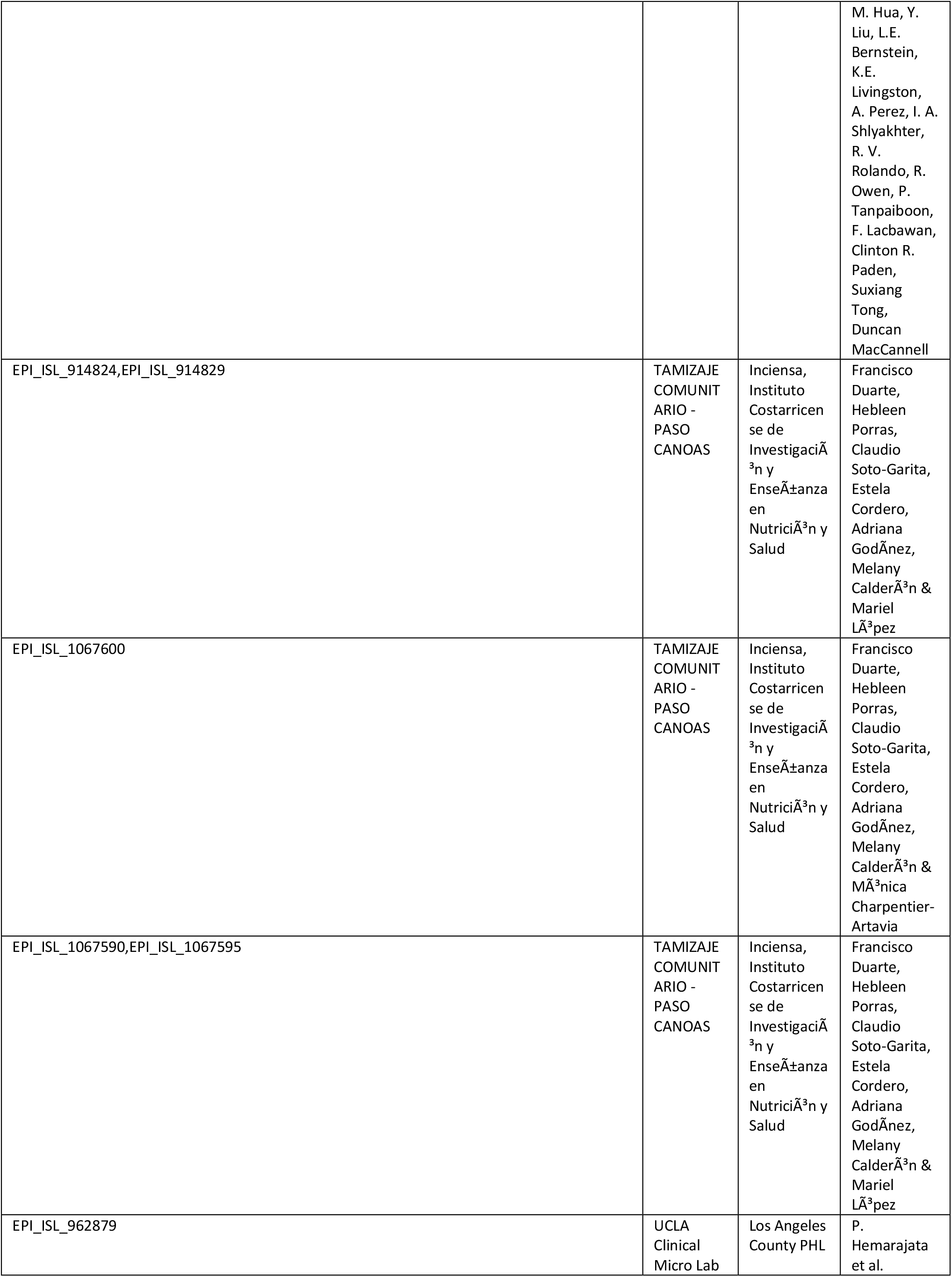
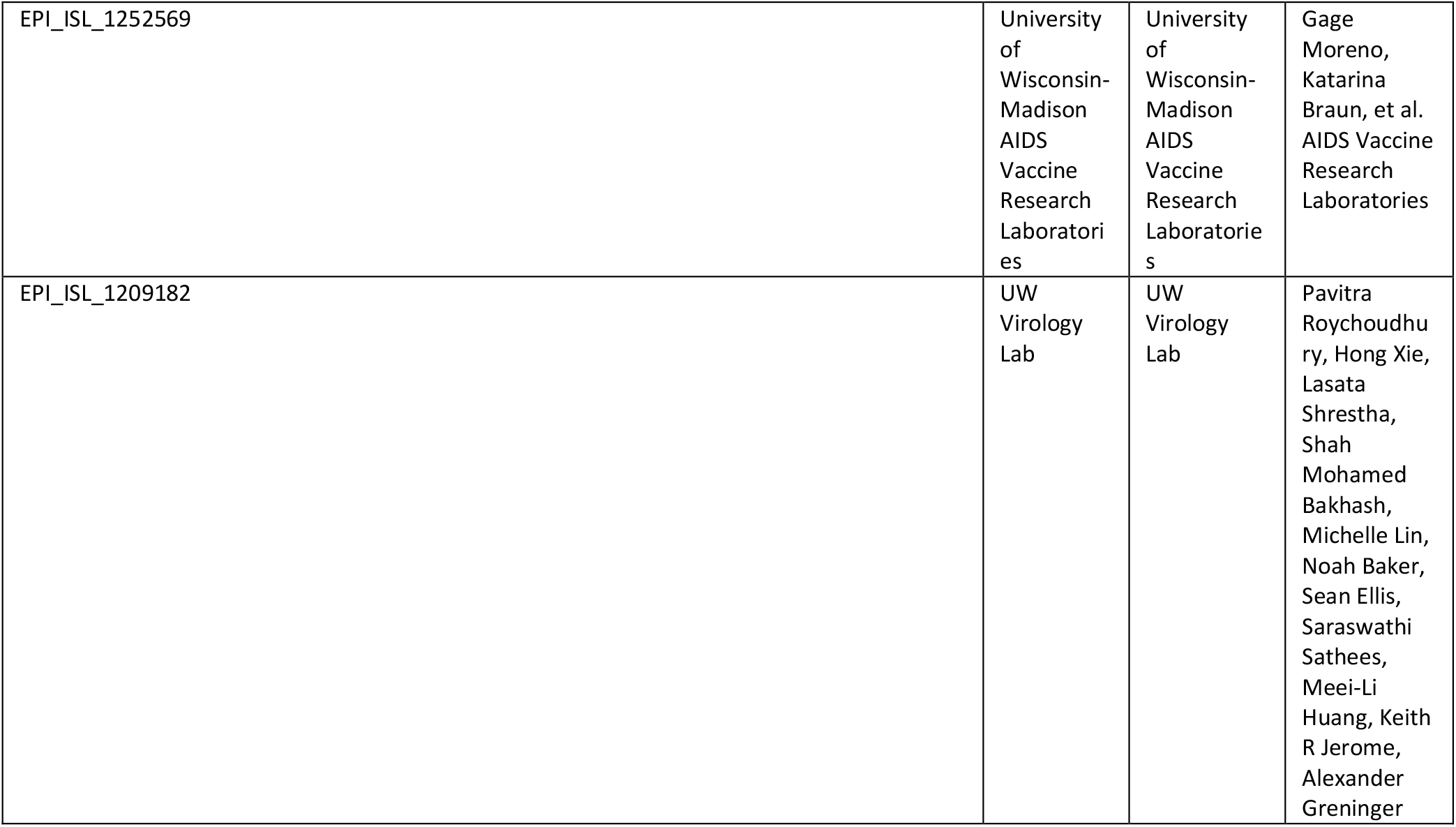
GISAID EpiCov Acknowledgments Table.

